# AT(N) predicts near-term development of Alzheimer’s disease symptoms in unimpaired older adults

**DOI:** 10.1101/2022.05.09.22274638

**Authors:** Cherie Strikwerda-Brown, Julie Gonneaud, Diana A. Hobbs, Frédéric St-Onge, Alexa Pichet Binette, Hazal Ozlen, Karine Provost, Jean-Paul Soucy, Rachel F. Buckley, Tammie L.S. Benzinger, John C. Morris, Victor L. Villemagne, Vincent Doré, Reisa A. Sperling, Keith A. Johnson, Christopher C. Rowe, Brian A. Gordon, Judes Poirier, John C.S. Breitner, Sylvia Villeneuve, the AIBL, Knight ADRC, HABS and PREVENT-AD research groups

## Abstract

**Importance:** National Institute on Aging-Alzheimer’s Association (NIA-AA) workgroups have proposed biological research criteria intended to identify individuals with preclinical Alzheimer’s disease (AD).

**Objective:** Assess the clinical value of these biological criteria for prediction of near-term cognitive impairment in cognitively unimpaired older individuals.

**Design, Setting, and Participants:** We studied 580 cognitively unimpaired older adults from four independent cohorts (PREVENT-AD: 128; HABS: 153; AIBL: 48; Knight ADRC: 251) having ≥1 year of clinical observation following Aβ and tau PET (median follow-up: PREVENT-AD = 3.16 yrs [1.51-4.50]; HABS = 1.94yrs [1.13-5.42]; AIBL = 3.66yrs [1.72-5.98]); Knight ADRC = 3.01 yrs [1.04-6.20]).

**Exposures:** Based on binary assessment of global amyloid burden (A) and of a composite temporal region of tau PET uptake (T), we stratified participants into four groups (A+T+, A+T-, A-T+, A-T-). Presence (+) or absence (-) of neurodegeneration (N) was assessed using temporal cortical thickness.

**Main Outcomes and Measures:** We analyzed each cohort separately. Primary outcome was clinical progression to mild cognitive impairment (MCI). A secondary outcome was cognitive decline. We compared MCI progression and cognitive decline across the four biomarker groups. MCI was identified by consensus committee review in PREVENT-AD, HABS, and AIBL, and by a CDR ≥ 0.5 in Knight ADRC. Clinical raters were blinded to imaging, genetic, and fluid biomarker data. Using a composite measure, cognitive decline was identified by a slope >1 SD below that of A-T- ‘non-progressors’.

**Results:** Across cohorts, 32 - 83% of A+T+ participants progressed to MCI during follow-up (mean progression time 2.0 - 2.72 years), as compared with <12% of participants in other biomarker groups. In two cohorts, progression increased to 100% when A+T+ individuals were also (N+). Cox proportional hazard ratios for progression to MCI in the A+T+ group vs. other biomarker groups were >5. Many A+T+ ‘non-progressors’ nonetheless showed longitudinal cognitive decline, while cognitive trajectories in other groups remained predominantly stable.

**Conclusions and Relevance:** Clinical prognostic value of the NIA-AA research criteria was confirmed in four independent cohorts, with nearly all A+T+(N+) cognitively unimpaired older individuals developing AD symptoms within ∼2-3 years.

**Key Points:** 

**Question:** What is the clinical relevance of the AT(N) biological classification of Alzheimer’s disease (AD) in unimpaired older adults?

**Findings:** In this prospective study of 580 cognitively unimpaired participants from four independent cohorts, between 31.58 and 100% of A+T+(N+) participants progressed to mild cognitive impairment (MCI) within 2-3 years after PET. The majority of A+T+ non-progressors also showed cognitive decline.

**Meaning:** Cognitively unimpaired older adults with biological AD are at imminent risk of developing MCI. These individuals may be ideal candidates for disease modifying therapies.

## Introduction

The National Institute on Aging-Alzheimer’s Association (NIA-AA) research criteria for Alzheimer’s disease (AD) were revised in 2018 to add tau biomarkers. In the resulting AT(N) framework, amyloid-beta (A) and tau (T) are needed for the diagnosis of AD, while neurodegeneration (N) is used to stage disease severity ^1^. These biomarkers can be classified as normal (-) and abnormal (+) such that individuals who are A+T+ can be said to have biological AD, even if they do not have cognitive symptoms. The clinical significance of biologically-defined AD in individuals without cognitive impairment remains debated ^2^, given that abnormal levels of amyloid-beta (Aβ) and tau are apparent in ∼20% of cognitively unimpaired older adults both *in vivo* ^3^ and at autopsy ^4^. As the cited studies are cross-sectional, however, it is unclear whether A+T+ individuals are at imminent risk of developing AD-related cognitive impairment. Regular near-term development of mild cognitive impairment (MCI) in cognitively unimpaired A+T+ individuals would provide strong evidence favoring a biological definition of pre-clinical AD. It would also have important implications both for clinical trial recruitment and prognosis of early clinical disease.

Using positron emission tomographic (PET) signal for Aβ or tau deposition across four independent cohorts, we investigated whether elevation of both biomarker signals predicted near-term progression from cognitively unimpaired to mild cognitive impairment (MCI). We also tested whether the evidence of neurodegeneration added clinical predictive value to the amyloid and tau PET biomarkers.

## Methods

### Participants and Study Design

Participants included 128 individuals from the family-history positive Pre-symptomatic Evaluation of Experimental or Novel Treatments for Alzheimer’s Disease (PREVENT-AD) cohort, 153 from the Harvard Aging Brain Study (HABS), 48 from the Australian Imaging, Biomarker & Lifestyle (AIBL) study, and 251 from the Knight Alzheimer Disease Research Center (ADRC) dataset (details in Supplement). All participants included in this study had at least one Aβ and tau PET scan, were cognitively unimpaired at the time of PET, and had at least 12 months of clinical follow-up thereafter. Participants provided written informed consent, and research procedures were approved by the relevant ethics committees. All analyses were performed separately for each cohort.

Full details of all measures, outcomes, their relative timing, and analyses are contained within the Supplement.

### Cognitive evaluation

All participants completed the Mini Mental State Examination (MMSE) ^5^ at the time of tau PET and had longitudinal cognitive testing using a composite measure specific to each cohort. The Repeatable Battery for the Assessment of Neuropsychological Status (RBANS) ^6^ was used in PREVENT-AD, and the Preclinical Alzheimer’s Cognitive Composite score (PACC) used in the other cohorts ^7,8^. Performance was evaluated using cohort-derived z-scores. Tau PET was introduced mid-study in all cohorts. All participants were required to be cognitively unimpaired both at cognitive baseline and at the time of PET.

### Outcomes

The primary outcome measure was clinical progression to MCI following PET among cognitively unimpaired participants. This outcome was adjudicated in all instances by persons masked to PET and MRI data, and to *APOE* genotype. In PREVENT-AD, HABS, and AIBL, MCI classifications were made by consensus committees comprising expert clinical and research staff. In the Knight ADRC, MCI was defined by a Clinical Dementia Rating® (CDR®)^9^ score of ≥0.5. Median follow-up after PET ranged from 1.94 to 3.66 years across cohorts. A secondary outcome was cognitive decline, as defined by a longitudinal slope in composite cognitive scores ≥ 1 SD below the mean of the A-T- non-progressors ^10,11^. For this outcome, we took advantage of the full length of study follow-up (including pre-PET) to create the slopes and characterise participants as “decliners” vs “non-decliners” (median follow-up across cohorts 5.10 - 6.31 yrs; minimum: 0.90 - 3.26 yrs; maximum: 7.26 - 14.47 yrs).

### A/T/(N) classification

Aβ PET imaging was performed using [^18^F]NAV4694 (NAV) in PREVENT-AD, [11C] Pittsburgh Compound B (PiB) in HABS, [^18^F]AV45 (florbetapir) and NAV in AIBL, and PiB and florbetapir in Knight ADRC (processing details in Supplement). Tau PET was performed using [^18^F]AV1451 (flortaucipir; FTP) in all cohorts ^12-16^. T1-weighted structural MRI scans were collected on 3T scanners and segmented with the Freesurfer Desikan-Killiany atlas ^17^. Pre- processing was performed using cohort-specific pipelines, and did not include partial volume correction. Standardized uptake value ratios (SUVRs) (distribution volume ratios (DVRs) for PIB) for each Desikan-Killiany region were computed using the cerebellum grey matter for all scans except for tau PET scans in PREVENT-AD, which used inferior cerebellar grey matter ^18^.

Participants were allocated to four PET biomarker groups (A+T+, A+T-, A-T+, A-T-). Cohort-specific thresholds were employed to establish Aβ positivity based on a global amyloid index^19^ (Centiloid values: PREVENT-AD = 22.32; HABS = 23.9; AIBL = 25; Knight ADRC = 27.1 and 21.9 for PiB and florbetapir, respectively; see Supplement for SUVR/DVR). A temporal meta-ROI was used as the primary measure of tau positivity. This comprised the average SUVR of the bilateral entorhinal cortex, amygdala, fusiform gyrus, and inferior and middle temporal gyri^20^. Tau positivity was defined as meta-ROI uptake surpassing 2 SD from the mean of cognitively unimpaired (at baseline) Aβ- participants in each cohort (SUVR cut-offs: PREVENT-AD = 1.30; HABS = 1.31; AIBL = 1.32; Knight ADRC = 1.28).

In secondary analyses, the presence (+) or absence (-) of neurodegeneration (N) was designated based on average cortical thickness in a bilateral temporal meta-ROI comprising entorhinal cortex, fusiform, inferior temporal, and middle temporal gyri ^17^. Participants were classified as neurodegeneration positive if temporal cortical thickness was below the 20^th^ percentile of A-T- non-progressor participants within the respective cohorts.

### Statistical Analysis

Analyses were performed separately for each cohort to assess replicability of results across samples and methodologies. The A-T+ group was excluded from statistical comparisons due to the low number of participants (PREVENT-AD: 1, HABS: 4; AIBL: 1; Knight ADRC: 4), though data from this group are presented visually for completeness. For demographic and clinical variables, we used one-way analyses of variance with Tukey’s post hoc tests to compare biomarker groups on continuous variables, and Fisher’s exact tests for categorical variables, including progression status. Cox proportional hazard models tested whether the risk of MCI progression over time was higher in the A+T+ group relative to the other PET biomarker groups, including age, sex, education, and *APOE* ε4 status as covariates. In follow-up analyses, continuous measures of neurodegeneration (temporal cortical thickness or hippocampal volume) were added to the PET biomarker Cox models. These were used instead of categorical AT(N) status given the very small sample size of each AT(N) group. We then compared the performance of each of these AT(N) models with clinical models that included MMSE, age, sex, education and *APOE* ε4 status. Of note, while progression to MCI was adjudicated blind to the AT(N) biomarkers, it was not blind to MMSE performance in the PREVENT-AD, HABS, or AIBL cohorts. Finally, we employed linear mixed-effects models with random slopes and intercepts to investigate longitudinal cognitive decline across the different AT(N) groups. This secondary outcome was intended to explore whether individuals who had not yet progressed to MCI were nonetheless likely to be on a clinical pathway toward AD symptoms. To do so, participants were further divided into cognitively ‘stable’ versus ‘decliners’ based on individual longitudinal cognitive slopes. The proportion of cognitive decliners versus cognitively stable in each biomarker group were then compared using Fisher’s exact tests.

We also performed sensitivity analyses in which analyses were repeated using 1) other commonly used regions to define tau PET positivity and 2) hippocampal volume to define neurodegeneration.

Alpha was set at *p* < .05 for all analyses. Analyses were performed using R Studio v1.1.463.

## Results

### Demographic and biological characteristics across biomarker groups

Across cohorts, between 7.19 and 12.50% of participants were classified as A+T+, compared with 20.83 to 24.22% as A+T-, 0.78 to 2.61% A-T+, and 64.58 to 68.13% A-T- (see Supplement for groupings using other regions to define T+). In PREVENT-AD, the A+T+ group included one participant originally classified as T- using the temporal meta-ROI quantification, but who displayed extensive occipital tau binding upon visual inspection (Figure 2, row 3). Characteristics of participants across cohorts and biomarker groups are presented in Table 1 (see Supplement for statistics, and characteristics by MCI progression status).

**Table 1.** Demographic, pathological and clinical characteristics of participants by biomarker group across cohorts.

|  | PREVENT-AD |  |  |  |  | HABS |  |  |  |  | AIBL |  |  |  |  | Knight ADRC |  |  |  |  |
| --- | --- | --- | --- | --- | --- | --- | --- | --- | --- | --- | --- | --- | --- | --- | --- | --- | --- | --- | --- | --- |
|  | Full sample<br>(n = 128) | A+T+<br>(n = 13) | A+T-<br>(n = 31) | A-T+<br>(n = 1) | A-T-<br>(n = 83) | Full sample<br>(n = 153) | A+T+<br>(n = 11) | A+T-<br>(n = 36) | A-T+<br>(n = 4) | A-T-<br>(n = 102) | Full sample<br>(n = 48) | A+T+<br>(n = 6) | A+T-<br>(n = 10) | A-T+<br>(n = 1) | A-T-<br>(n = 31) | Full sample<br>(n = 251) | A+T+<br>(n = 19) | A+T-<br>(n = 57) | A-T+<br>(n = 4) | A-T-<br>(n = 171) |
| **Demographics** |  |  |  |  |  |  |  |  |  |  |  |  |  |  |  |  |  |  |  |  |
| Age, years | 67.35<br>(4.87) | 71.32<br>(5.12)<br><sub>ab</sub> | 66.72<br>(4.57) | 60-65 | 67.00<br>(4.73) | 76.11<br>(6.33) | 78.48<br>(5.21) | 77.47<br>(6.17) | 84.06<br>(3.72) | 75.06<br>(6.26) | 74.71<br>(6.87) | 79.17<br>(6.55)<sup>b</sup> | 79.50<br>(7.76)<sup>c</sup> | 80-85 | 72.13<br>(5.40) | 71.97<br>(5.73) | 75.02<br>(5.81)<sup>b</sup> | 72.10<br>(5.38) | 70.79<br>(4.74) | 71.62<br>(5.79) |
| Sex, F:M<br>(% F) | 95:33<br>(74.22) | 10:3<br>(76.92) | 25:6<br>(80.65) | 0:1<br>(0) | 60:23<br>(72.29) | 86:67<br>(56.21) | 9:2<br>(81.82) | 18:18<br>(50) | 2:2<br>(50) | 57:45<br>(55.88) | 29:19<br>(60.42) | 5:1<br>(83.33) | 6:4<br>(60) | 1:0<br>(100) | 17:14<br>(54.84) | 137:114<br>(54.58) | 15:4<br>(78.95)<sup>b</sup> | 36:21<br>(63.16) | 3:1<br>(75) | 83:88<br>(48.54) |
| Education, years | 15.17<br>(3.28) | 13.46<br>(2.73) | 15.19<br>(3.00) | 20<br>(0) | 15.37<br>(3.39) | 16.08<br>(3.06) | 17.09<br>(2.07) | 16.06<br>(2.87) | 18<br>(1.63) | 15.91<br>(3.23) | 11.79<br>(2.97) | 9.67<br>(2.73) | 12.20<br>(2.86) | 15<br>(0) | 11.97<br>(2.96) | 16.33<br>(2.38) | 15.95<br>(2.30) | 16.63<br>(2.24) | 16.00<br>(1.63) | 16.28<br>(2.45) |
| APOE ε4 carriers, n (%) | 50<br>(39.06) | 9<br>(69.23)<br><sub>b</sub> | 18<br>(58.06)<br><sub>c</sub> | 0 (0) | 23<br>(27.71) | 46<br>(30.07) | 10<br>(90.91)<br><sub>ab</sub> | 20<br>(55.56)<br><sub>c</sub> | 1<br>(25) | 15<br>(14.71) | 12 (25) | 2<br>(33.33) | 3 (30) | 0 (0) | 7<br>(22.58) | 76<br>(30.28) | 13<br>(68.42)<sup>ab</sup> | 26<br>(45.61)<sup>c</sup> | 0 (0) | 37<br>(21.64) |
| **PET** |  |  |  |  |  |  |  |  |  |  |  |  |  |  |  |  |  |  |  |  |
| Global Aβ Centiloid | 26.59<br>(28.76) | 75.87<br>(40.36)<br><sub>ab</sub> | 44.92<br>(28.24)<br><sub>c</sub> | 18.33 | 12.13<br>(5.12) | 19.00<br>(20.52) | 52.86<br>(12.85)<br><sub>ab</sub> | 43.29<br>(18.60)<br><sub>c</sub> | 10.45<br>(1.42) | 7.11<br>(4.05) | 19.35<br>(38.44) | 72.33<br>(34.33)<br><sub>a</sub> | 64.10<br>(23.26)<br><sub>c</sub> | -16 | -4.19<br>(10.60) | 21.03<br>(33.51) | 79.81<br>(37.05)<sup>ab</sup> | 55.46<br>(30.16)<sup>c</sup> | 6.67<br>(7.95) | 3.35<br>(10.09) |
| Temporal meta-ROI SUVR | 1.19<br>(0.12) | 1.42<br>(0.18)<br><sub>ab</sub> | 1.18<br>(0.06) | 1.30 | 1.15<br>(0.07) | 1.19<br>(0.09) | 1.41<br>(0.05)<br><sub>ab</sub> | 1.20<br>(0.07)<br><sub>c</sub> | 1.33<br>(0.01) | 1.16<br>(0.06) | 1.19<br>(0.16) | 1.51<br>(0.16)<br><sub>ab</sub> | 1.18<br>(0.10) | 1.33 | 1.12<br>(0.09) | 1.16<br>(0.09) | 1.35<br>(0.09)<sup>ab</sup> | 1.16<br>(0.07)<sup>c</sup> | 1.30<br>(0.02) | 1.13<br>(0.07) |
| **MRI** |  |  |  |  |  |  |  |  |  |  |  |  |  |  |  |  |  |  |  |  |
| Temporal cortical thickness | 2.89<br>(0.11) | 2.83<br>(0.12)<sup>a</sup> | 2.95<br>(0.09)<br><sub>c</sub> | 2.91 | 2.88<br>(0.12) | 2.86<br>(0.16) | 2.72<br>(0.19)<sup>b</sup> | 2.84<br>(0.18) | 2.88<br>(0.13) | 2.88<br>(0.14) | 2.88<br>(0.11) | 2.82<br>(0.09) | 2.86<br>(0.11) | 3.02 | 2.90<br>(0.11) | 2.84<br>(0.14) | 2.81<br>(0.17) | 2.87<br>(0.13) | 2.82<br>(0.15) | 2.83<br>(0.14) |
| Hippocampal volume (% of TIV) | 0.54<br>(0.06) | 0.52<br>(0.06) | 0.57<br>(0.06) | 0.56 | 0.54<br>(0.06) | 0.48<br>(0.06) | 0.45<br>(0.06)<sup>b</sup> | 0.47<br>(0.05) | 0.46<br>(0.04) | 0.49<br>(0.06) | 0.51<br>(0.06) | 0.50<br>(0.02) | 0.48<br>(0.04) | 0.57 | 0.52<br>(0.06) | 0.51<br>(0.07) | 0.47<br>(0.07)<sup>ab</sup> | 0.52<br>(0.07) | 0.51<br>(0.07) | 0.51<br>(0.07) |
| **Cognition** |  |  |  |  |  |  |  |  |  |  |  |  |  |  |  |  |  |  |  |  |
| MMSE (/30) | 28.80<br>(1.26) | 27.85<br>(1.57)<br><sub>ab</sub> | 29.19<br>(0.79) | 30 | 28.78<br>(1.29) | 29.26<br>(0.98) | 28.45<br>(1.21)<sup>ab</sup> | 29.31<br>(0.86) | 29.75<br>(0.50) | 29.31<br>(0.97) | 28.46<br>(1.61) | 25.83<br>(2.04)<sup>ab</sup> | 28.80<br>(1.48) | 27 | 28.90<br>(1.01) | 29.27<br>(1.08) | 29.16<br>(1.17) | 29.35<br>(1.04) | 29.00<br>(1.41) | 29.27<br>(1.09) |
| RBANS, baseline |  |  |  |  |  |  |  |  |  |  |  |  |  |  |  |  |  |  |  |  |
| Global Cognition | -0.09<br>(0.88) | -0.36<br>(0.98) | 0.00<br>(0.89) | 0.32 | -0.09<br>(0.87) | NA | NA | NA | NA | NA |  |  |  |  |  |  |  |  |  |  |
| PACC, baseline | NA | NA | NA | NA | NA | 0.11<br>(0.61) | 0.13<br>(0.55) | 0.11<br>(0.55) | -0.02<br>(0.25) | 0.11<br>(0.66) | -0.46<br>(0.89) | -0.59<br>(0.58) | -0.71<br>(0.95) | -1.40 | -0.33<br>(0.92) | 0.00<br>(0.69) | -0.14<br>(0.58) | 0.04<br>(0.58) | 0.15<br>(0.35) | 0.00<br>(0.75) |

Data are presented as mean (standard deviation) unless otherwise specified. Aβ = amyloid beta; APOE = apolipoprotein E genetic
locus; DVR = distribution volume ratio; meta-ROI = meta region-of-interest; MCI = Mild Cognitive Impairment; MMSE = Mini
Mental State Examination; PACC = Preclinical Alzheimer's Composite Score; PET = Positron Emission Tomography; RBANS =
Repeatable Battery for the Assessment of Neuropsychological Status; SUVR = standardized uptake value ratio; TIV = total
intracranial volume.
*Notes:* Age and MMSE performance were calculated at the time of tau PET. Education data was collected in ranges in AIBL, with the lower boundary of the range used in current analyses. Years of education are therefore likely underestimated in this cohort (further details in Supplement). *APOE* ε4 carriers had at least one copy of the ε4 allele. Cognitive variables (RBANS, PACC) are reported as cohort-derived z-scores. <sup>a</sup> = significant difference between A+T+ and A+T- groups, <sup>b</sup> = significant difference between A+T+ and A-T- groups, <sup>c</sup> = significant difference between A+T- and A-T- groups at $p < .05$ . The A-T+ group is presented for completion but was not included in statistical analysis owing to its small sample size.

### Clinical progression rates across biomarker groups

Between 6.54% and 16.67% of participants across cohorts progressed from cognitively unimpaired to MCI after PET (mean progression time: PREVENT-AD = 2.00 years (SD = 1.10); HABS = 2.72 years (SD = 1.49), AIBL = 2.55 years (SD = 1.18), Knight ADRC = 2.67 years (SD = 1.18)). MCI progression status by biomarker group is displayed in Figure 1 and Supplementary Table 2. Examples of Aβ and tau PET scans for each biomarker group and cognitive status from PREVENT-AD are presented in Figure 2. Across all cohorts, a greater proportion of A+T+ participants progressed to MCI (ADRC: 31.58%, HABS: 45.45%, PREVENT-AD: 61.54%, AIBL: 83.33%) compared with the other PET biomarker groups (<20%) (*p* values ≤ .001; Figure 1A-D & Supplement). Compared with other regions (entorhinal, inferior temporal, or ‘any’), the meta-ROI analysis for tau positivity detected the highest proportion of MCI progressors in the PREVENT-AD, HABS and Knight ADRC cohorts, whereas an inferior temporal ROI detected the highest proportion of MCI progressors in AIBL (100% vs 83.33%) (Supplement). In A+T+ participants, evidence of neurodegeneration (N+), defined using temporal cortical thickness, was associated with a 37.50 to 100% MCI progression rate (Figure 1E-H & Supplement). Results were similar using hippocampal volume (Supplement).

**Figure 1.**
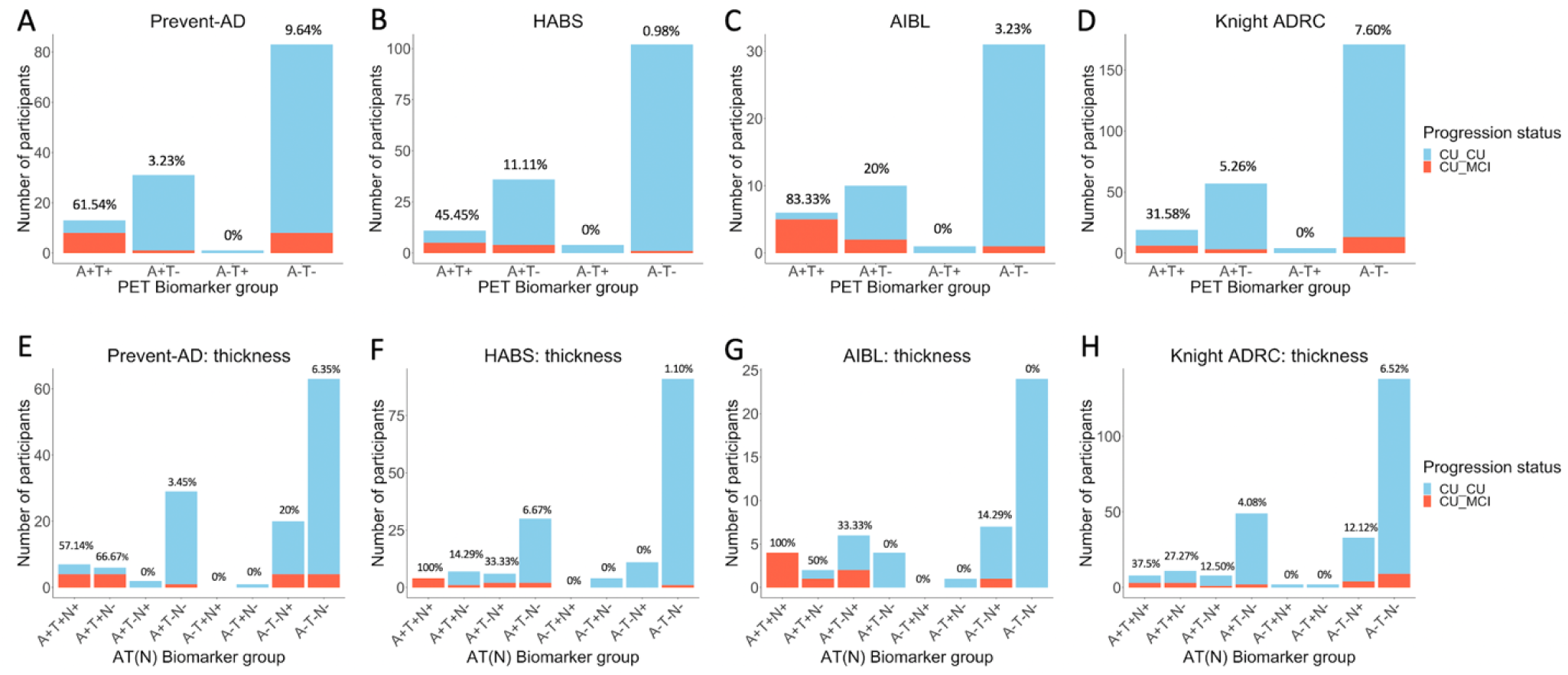
Number of participants progressing to MCI after PET versus those remaining cognitively unimpaired in each AT (Panels A-D) and AT(N) (Panels E-H) biomarker group, across cohorts. Percentage values represent the proportion of MCI progressors within the group. CU_CU = Cognitively unimpaired at time of PET, remaining cognitively unimpaired during follow-up; CU_MCI = Cognitively unimpaired at time of PET, progressing to MCI during follow-up. *Note:* The A-T+ group is displayed for visualisation purposes but was not included in statistical analysis due to the small sample size. While the MCI classifications were based on clinical consensus in the PREVENT-AD, HABS and AIBL cohorts, these were based on a CDR of ≥0.5 for the Knight ADRC cohort. (N) was defined by temporal cortical thickness.

**Figure 2.**
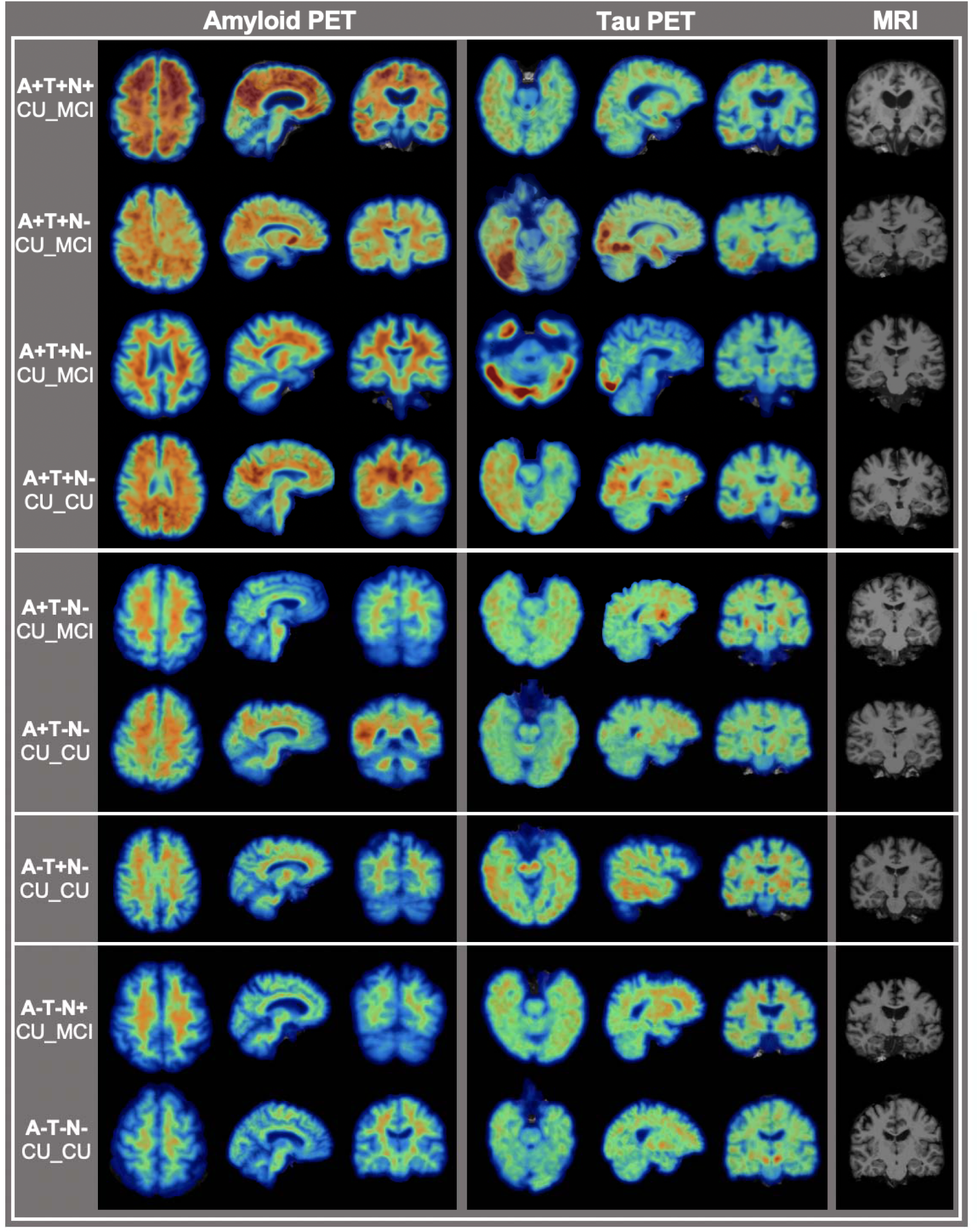
Example Aβ and tau PET and MRI scans from different AT(N) biomarker groups in the PREVENT-AD cohort. Of note, the subject in row 3 was initially classified as T- using the temporal meta-ROI quantification, but upon visual inspection was found to harbour significant tau burden in the occipital lobe and was therefore reclassified as T+. Demographics of the participants, in descending order, are as follows: female 70-75y; male 65-70y; female 70-75y; female 60-65y; female 60-64y, female 65-70y; male 60-65y; female 60-65y; female 65-70y. CU_CU = Cognitively unimpaired at time of PET, remaining cognitively unimpaired during follow-up; CU_MCI = Cognitively unimpaired at time of PET, progressing to mild cognitive impairment during follow-up.

### Effect of biomarker group on probability of clinical progression across time

Survival curves representing progression time from CU to MCI for each AT biomarker group are displayed in Figure 3. Given shorter follow-ups in the A+T+ group compared with the other biomarker groups in HABS, data for all participants in this cohort were censored at the last available time point within the A+T+ group (2.59 years; see Supplement for models including all data). Using the meta-ROI to define T+, the A+T+ group demonstrated a greater probability of progression to MCI over time compared with the other groups (Hazard Ratios > 5.79, Model Likelihood Ratios > 19.37, *p* values < .005, Concordance > 0.76; Figure 3 & Supplement). Biomarker group hazard ratios were typically reduced in magnitude when other regions were used to define tau positivity (Supplement). Models including the AT biomarker groups outperformed those using demographic/clinical information alone (Model Likelihood Ratios < 27.04; Supplement). Continuous measurement of neurodegeneration did not add significant predictive value for MCI progression in the biomarker group models (*p* values > .09), with the exception of cortical thickness in HABS and Knight ADRC (*p* values = .03 and .04, respectively) (Supplement).

**Figure 3.**
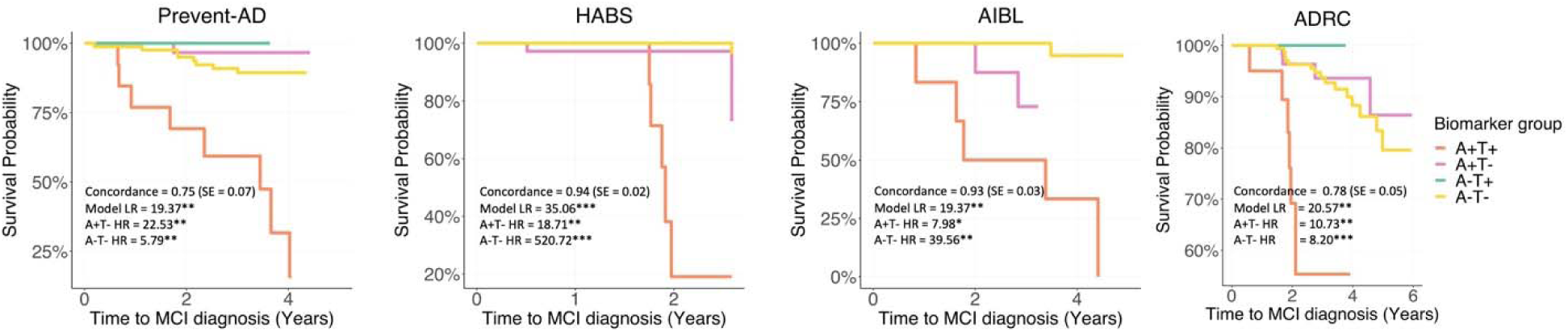
(A-D) Survival curves reflecting time from PET scan to MCI classification for the four biomarker groups, across cohorts, using the temporal meta-ROI to define T+. Model likelihood ratios (LR), concordance values and standard errors (SE) reported for Cox regression models included age, sex, education and *APOE* ε4 status at the time of tau PET as covariates. Hazard ratios (HR) for the PET biomarker groups are in reference to the A+T+ group. Inverted hazard ratios are reported for ease of interpretation (i.e., reflecting risk of progression to MCI rather than survival i.e., non-progression). *Notes:* Data was censored at the date of MCI classification (for progressors), or the last clinical follow-up visit for each participant (for non-progressors), with the exception of the HABS cohort for which data was censored at the date of MCI classification (for progressors), or the last available time point within the A+T+ group (i.e., 2.59 years) for non-progressors, given uneven follow-up times between the biomarker groups. The A-T+ group is displayed for visualisation purposes but was not included in statistical analysis due to the small sample size. * = *p* < .05, ** = *p* < .01, *** *p* < .001.

### Longitudinal cognition across biomarker groups

In all cohorts, A+T+ participants experienced greater longitudinal cognitive decline compared with the other groups (all β estimates > 0.04, *p* values < .03; Supplement). The strength of these associations was reduced when other regions were used to define tau abnormality in PREVENT-AD and HABS, whereas the inferior temporal lobe performed better in AIBL and Knight ADRC (Supplement). Figure 4A displays longitudinal cognitive performance for each PET-biomarker group (see Supplement for performance stratified by MCI progression status and for specific RBANS cognitive indexes in PREVENT-AD).

**Figure 4.**
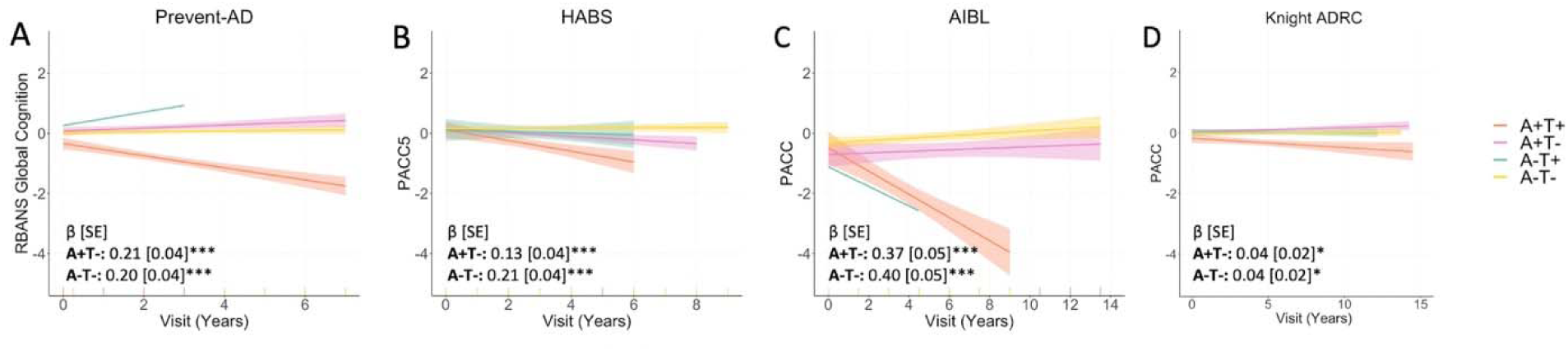
(A-D) Longitudinal cognitive slopes for each biomarker group across cohorts. Models included random slopes and intercepts for each subject and covariates of age, sex, and years of education. *Notes:* For all cohorts PET was added mid-study, and was therefore performed at different cognitive follow-up visits for each participant. Longitudinal cognition analyses included time points both prior to and after PET scanning. The A-T+ group is displayed for visualisation purposes but was not included in statistical analysis due to the small sample size. β estimates are for the interaction between biomarker group and visit date, with A+T+ as the reference group. Cognitive scores are reported as cohort-derived z-scores. * = *p* < .05, *** = *p* < .001.

### Cognitive decline status of non-progressors across biomarker groups

Cognitive status (declining versus stable) for non-progressors by biomarker group is displayed in Supplementary Figure 10. In PREVENT-AD and Knight ADRC, a greater proportion of A+T+ non-progressors showed cognitive decline (80% & 30.77%, respectively) compared with the A+T- and A-T- groups (≤16% in PREVENT-AD, ≤9.50 in Knight ADRC, *p* values < .05, Supplement). No group difference reached significance in the other cohorts, but adding the decliners to the progressors in AIBL captured 100% of the A+T+ participants (Supplement). Using different regions to classify tau positivity produced varying results across cohorts, with regions other than the temporal meta-ROI performing better at capturing decliners in some cases (Supplement).

## Discussion

The AT(N) biological framework for AD has been proposed for research purposes, ^1^ but its clinical significance for individuals without cognitive impairment is unclear. We examined the implications of Aβ and tau positive PET signals for clinical progression from cognitively unimpaired to MCI over short-term intervals. Across four independent cohorts, 32% - 83% of cognitively unimpaired individuals with abnormal elevation of both Aβ and tau progressed to MCI within a mean of 2 to 2.7 years after PET scanning. These numbers increased in three out of the four cohorts when restricted to (N+) individuals, reaching 100% progression rate in two of them. Most of the remaining A+T+ participants also experienced cognitive decline, suggesting that they too are on a pathway towards AD symptoms.

AD clinical trials often require an abnormal amyloid biomarker for inclusion ^21,22^. Here, positivity on both Aβ and tau PET was associated with an 8 to 23 times greater hazard of progression from cognitively unimpaired to MCI, as compared with a positive Aβ scan in the absence of a tau-positivity (A-T+ group results not considered, given this group represented <3% of all participants). These results suggest 1) that the presence of Aβ is typically needed as a precondition to tau-PET tracer binding detection, and 2) that tau pathology is critical for imminent decline. Models based on A+T+ PET biomarkers significantly outperformed models based on demographic and clinical data alone as predictors of progression to MCI. Combining both tau and Aβ PET therefore greatly boosts the near-term prognostic prediction of clinical progression in preclinical disease stages – a finding that is highly relevant for future clinical trials. Examination of longitudinal cognitive trajectories further indicated that 31% - 100% of A+T+ participants who remained ‘cognitively unimpaired’ nonetheless demonstrated significant cognitive decline. The vast majority of individuals with neither Aβ nor tau pathology (A-T-) maintained stable cognition over time, regardless of their clinical classification.

The research framework for the biological definition of AD uses dichotomous categories to define biomarker abnormality, i.e., (+) or (-)^1^. One challenge for this framework in PET studies is choice of anatomical region from which to define tau positivity ^23^. While the entorhinal cortex (EC) is often the site of earliest tau deposition in AD ^24^, tau in this region is not necessarily specific to AD and may also occur with increasing age, independent of Aβ ^25,26^. Accordingly, a temporal meta-ROI, comprising both medial and neocortical temporal regions, has been proposed as an alternative to the EC ROI for detecting AD-specific early tau deposition ^19^. Here, use of a temporal meta-ROI to define T+ identified a larger percentage of MCI progressors in the A+T+ group and showed stronger associations with longitudinal cognitive decline when compared to the EC ROI. However, in one of the four cohorts investigated here, an inferior temporal tau ROI slightly surpassed the temporal meta-ROI in detecting A+T+ MCI progressors. This cohort represented only a fraction (48/580) of all participants.

Evidence of neurodegeneration is not required for a diagnosis of biological AD, but is thought instead to reflect a non-specific marker of disease severity typical of more advanced stages. In the HABS cohort, A+T+ individuals with thinner temporal cortices had increased MCI progression rates. In the other cohorts, the progression rate was not significantly higher in the A+T+(N+) than the A+T+(N-) group when cortical thickness was used to define (N+), though it is notable that 100% of the A+T+(N+) AIBL participants progressed to MCI. The percentage of MCI progression also increased from 50% to 80% in the PREVENT-AD A+T+(N-) and A+T+(N+) groups respectively when (N) was defined based on the hippocampal volume. The absence of a significant difference in AIBL, PREVENT-AD, and Knight ADRC may be attributable to the very high percentage of A+T+ progressors in AIBL (83%) and low statistical power in PREVENT-AD and Knight ADRC.

A key strength of our study was the replicability of results across four independent cohorts using related but different methods, as well as the robustness of the reported findings in multiple sensitivity analyses. Study limitations include the relatively modest sample sizes of the A+T+ groups, though the proportion of participants assigned to this biomarker group was similar to previous studies ^27-29^. The maximum duration of clinical follow-up post-PET also varied across individuals, and was overall quite short (median = 1.94 - 3.66 years). Altogether these results suggest that individuals with both amyloid and tau do not remain cognitively normal for a long period of time. Among A+T participants who do remain in a “cognitively normal” category, the common appearance of substantial longitudinal decline in cognition suggests the possibility of subsequent progression to MCI.

## Conclusions

In four independent cohorts, we demonstrate that Aβ and tau PET positivity in cognitively unimpaired individuals is a substantial predictor both of near-term progression to MCI and, among those who do not show such categorical change, of longitudinal cognitive decline. Additional evidence of neurodegeneration (N) implies substantial additional probability of clinical progression. Crucially, abnormality in both Aβ and tau PET was associated with a considerably greater risk of near-term clinical progression than abnormality of Aβ PET alone.

These findings support the clinical validity of a biological definition of AD in cognitively unimpaired subjects. When preventive treatments become available, the use of such a biological definition of AD to identify persons with probable pre-clinical AD could substantially mitigate the AD epidemic. Until then, elevations in both Aβ and tau PET indicate imminent clinical progression in cognitively unimpaired individuals.

## Data Availability

All data produced in the present study are available upon reasonable request to the authors

## Acknowledgment

CSB is supported by a joint Postdoctoral Fellowship from Canadian Institutes of Health Research (CIHR), Alzheimer’s Society of Canada (ASC), and Fonds de recherche du Québec – Santé. FS is supported by the Canada First Research Excellence Fund and Fonds de recherche du Québec, awarded to the Healthy Brains, Healthy Lives initiative at McGill University. APB is supported by a Brightfocus Foundation Postdoctoral Fellowship. JPS receives fees from Biogen Canada and is involved in pro bono projects with Optina Diagnostics. RFB is supported by a K99/R00 award from the National Institute of Aging (R00AG061238-03) and an Alzheimer’s Association Research Fellowship (AARF- 20-675646). JCM is supported by the National Institute of Health (NIH). VLV reports grants from Life Molecular Imaging, Avid Radiopharmaceuticals, and General Electric Healthcare. RAS and KAJ are involved in public-private partnership clinical trials sponsored by the NIH and Eli Lilly, which owns the distribution rights to flortaucipir, but they do not have any personal financial relationship with Eli Lilly. CCR has received speaker fees from Nutricia, Biogen and Roche, Scientific Advisory Board fees from Cerveau Technology and Prothena, payment for preparation of educational materials from Biogen and grants to his institution from Cerveau Technologies, Biogen, Abbvie, Actinogen and Eisai and Australian Government support from NHMRC grants APP1011689, APP10475151, APP1071430. BAG is supported by the Alzheimer’s Association and NIH. SV is supported by a Canada Research Chair and a Canada Fund for Innovation grant. The PREVENT-AD was funded by CIHR, Alzheimer’s Association, ASC, Brain Canada, McGill University, the government of Canada, an unrestricted gift from Pfizer Canada, the Canada Fund for Innovation, the Douglas Hospital Research Centre, and the Levesque Foundation, and Genome Quebec Innovation Center. Funding sources had no role in the design and conduct of the study; collection, management, analysis and interpretation of the data; preparation, review, or approval of the manuscript; and the decision to submit the manuscript for publication.

