## Supplementary for "AT(N) predicts near-term development of Alzheimer’s disease symptoms in unimpaired older adults"

Sylvia Villeneuve

Douglas Mental Health University Institute

Perry Pavilion Room E3417.1, 6875 Boulevard LaSalle

Montreal, QC, Canada H4H 1R3

**Supplementary Methods**

*Participants and Study Design*

*PREVENT-AD*

One-hundred twenty-eight participants were included from the Presymptomatic Evaluation of Experimental or Novel Treatments for Alzheimer’s Disease (PREVENT-AD) cohort, a longitudinal observational study of older adults at risk for sporadic AD ^1^. Enrolment criteria included 1) having a parent or at least 2 siblings with a history of AD; 2) age > 60 years (or between 55 and 59 if they were within 15 years of their youngest affected relative’s symptom onset); 3) no major neurological or psychiatric diseases; and 4) no cognitive impairment at study enrolment ^2^.

*HABS*

One-hundred fifty-three participants were taken from the Harvard Aging Brain Study (HABS), a longitudinal study of preclinical AD ^3^. For inclusion at study baseline, participants were required to score: 1) 0 on the Clinical Dementia Rating scale; 2) 11 or less on the Geriatric Depression Scale; 3) 27 or more on the education-adjusted Mini-Mental State Examination, and 4) within education-adjusted norms on Logical Memory – Delayed Recall.

*AIBL*

Forty-eight participants were included from the Australian Imaging, Biomarker & Lifestyle (AIBL) study^4^. Participants were required to be aged at least 60 years old, cognitively normal at study baseline as determined by clinical review (see further below), and be free from any major neurological or psychiatric history.

*Knight ADRC*

Two-hundred fifty-one participants were included from ongoing studies of aging and Alzhiemer’s Disease at the Charles F. and Joanne Knight Alzheimer Disease Research Center (ADRC), including the Adult Children Atudy (ACS) and the Health Aging and Senile Dementia (HASD) cohorts. Participants were greater than 60 years of age, and were classified as cognitively unimpaired with a CDR® score of 0 at study enrolment.

All studies were approved by the relevant ethics committees PREVENT-AD: Institutional Review Board of McGill University; HABS: Partners Healthcare Institutional Review Board; AIBL: Austin Health, St-Vincent’s Health, Hollywood Private Hospital and Edith Cowan University; Knight ADRC: Washington University in St. Louis Institutional Review Board).

*Measures*

*Relative timing*

Study enrolment for the PREVENT-AD study spanned the period from 2012 to 2017, while Aβ- and tau-PET scans were collected on consecutive days for each participant during 2017-2019. Cognitive assessments were performed approximately annually. PET was performed at different cognitive time points for each participant (baseline: 6; 1-year follow-up: 17; 2-year follow-up: 18; 3-year follow-up: 21; 4-year follow-up: 36; 5-year follow-up: 30 participants). For analyses focused on prediction of MCI progression based on PET, only cognitive assessments after the PET scan were included (median follow-up after PET scans = 3.21, mean = 3.16, range = 1.51-4.50 years), while for longtiduinal cognition analyses, the full length of study follow-up was used (median total follow-up = 5.44, mean = 5.22, range = 2.00-7.26 years).

Tau-PET imaging was introduced to HABS mid-study, with most participants completing tau-PET in Year 4 of study enrolment (n = 107). Cognitive assessments were administered approximately annually. A smaller number of participants underwent tau-PET at an earlier or later visit (Year 1: 3; Year 2: 16; Year 3: 19; Year 5: 8 participants). The Aβ-PET scan closest to the tau-PET scan was used for analyses (median days between = 93 days, range = 0-842). Median cognitive follow-up after PET was 1.94 years (mean = 2.35, range = 1.13-5.42), and median total follow-up was 5.10 years (mean = 5.19, range = 2.78-8.68).

In AIBL, study enrolment commenced in 2006, while tau-PET commenced in 2013. Cognitive measures were collected every 18 months. PET was performed at different cognitive time points for each participant (baseline: 6; 18-month follow-up: 16; 36-month follow-up: 8; 54-month follow-up: 2; 90-month follow-up: 15; 108-month follow-up: 1 participant). The closest Aβ-PET scan to the tau-PET scan was used for analyses (median days between = 302.50 days, range = 13-882). Median cognitive follow-up after PET was 3.66 years (mean = 3.96, range = 1.72-5.98), and median total follow-up was 6.31 years (mean = 7.78, range = 3.26-13.59).

Study enrolment for the Knight ADRC began in 1979 with the addition of tau-PET imaging in 2014. Aβ-PET scans collected nearest to the tau-PET scan was used for analyses (median days between = 28 days, range =1-523). Cognitive assessments (PACC) were collected on a roughly annual basis, though these did not always occur during the same visits as the clinical assessments (CDR). Accordingly, two participants completed tau-PET and at least 12 months of clinical follow-up, but only one cognitive assessment. These two participants were excluded from the longitudinal cognition analyses (one A+T+ and one A+T- participant). As tau-PET is a recent addition to the study, it was completed at various timepoints relative to participant enrolment (Baseline: 84, Year1: 27, Year2: 15, Year3: 35, Year4: 14, Year5: 25, Year6: 13, Year7: 20, Year8: 12, Year9: 2, Year10: 3, Year11: 1). Median cognitive follow-up after PET was 3.01 years (mean = 3.28, range = 1.05-6.20), and median total follow-up was 6.26 years (mean = 6.42, range = 0.90-14.47).

*Years of education*

In AIBL, years of education was recorded in ranges: < 7 years, 7-8 years, 9-12 years, 13-15 years, 15+ years. As a conservative estimate, we years of education in AIBL as the lower bound of the ranges, though acknowledge these are likely to represent an underestimate.

*Cognitive evaluation*

In PREVENT-AD, the Repeatable Battery for the Assessment of Neuropsychological Status (RBANS) ^5^ was administered annually. The RBANS comprises 12 cognitive tasks, which produce five cognitive domain Index scores (Immediate Memory, Attention, Visuospatial Construction, Language, and Delayed Memory), along with a total score measuring Global Cognition. Global Cognition was employed as the primary measure of interest, with results for the Index scores included in Supplementary Results.

In HABS, AIBL, and Knight ADRC the Preclinical Alzheimer’s Cognitive Composite (PACC) score was used to measure cognition ^6,7^. This composite measure assesses cognitive domains of memory, executive function, and semantic processing, though the exact tests of these processes differ between the two cohorts ^8^.

Cognitive scores represent z-scores calculated using cohort-specific means and standard deviations.

*Neuroimaging Acquisition*

PET imaging was performed in PREVENT-AD using [18F]NAV4694 (NAV) to assess Aβ burden and [^18^F]AV1451 (flortaucipir; FTP) for tau deposition. Aβ scans were acquired 40 to 70 minutes post-tracer injection (≈222 MBq), and tau scans 80 to 100 minutes post-injection (≈370 MBq). PET images were acquired at the McConnell Brain Imaging Centre at the Montreal Neurological Institute, generally across two consecutive days. T1-weighted structural MRI scans were acquired on a 3T Siemens Trio scanner at the Brain Imaging Centre of the Douglas Mental Health University Institute, with the following parameters: repetition time = 2300ms, echo time = 2.98ms, 176 slices, slice thickness = 1mm ^1^; median time between MRI and tau-PET scans: 275 days, range: 138-1008.

Acquisition and processing details of the HABS PET data have been described previously ^9,10^. Briefly, Aβ-PET scans were acquired using PiB (^11^C Pittsburgh Compound B) during a 60-minute dynamic acquisition beginning immediately after injection. FTP scans were acquired 80-100 minutes following a 9.0 to 11.0 mCi bolus injection. MRI scanning was performed on a 3T Tim Trio scanner with a 12-channel phased-array head coil. Imaging measures were typically collected every 2 years (median time between MRI and tau-PET scans: 106 days, range: 2-1221).

Imaging acquisition and preprocessing details for AIBL are described in ^11-13^. Aβ-PET scans were acquired using [^18^F]AV45 (florbetapir) and NAV, both involving a 20-min acquisition performed 50 min post-injection. FTP scans involved four 5-minute frames, commencing either 75-105 or 80-100 minutes post-injection. For structural MRI, high-resolution T1-weighted anatomical magnetization-prepared rapid gradient echo (MPRAGE) sequences were obtained using a Siemens 3T TIM Trio scanner (median time between MRI and PET scans: 312.50 days, range: 0-1467).

Detailed image acquisition and preprocessing methods for the Knight ADRC can be found in ^14,15^. Aβ-PET scans utilized PiB (30-60 min post-injection window) and [^18^F]AV45 (50-70 min post-injection window) to assess amyloid burden. Tau deposition was measured using [^18^F]AV1451 PET imaging acquired 80-100 min following a 7.2 to 10.8 mCi injection. High-resolution T1-weighted MPRAGE images were collected on 3T Siemens Biograph mMR PET-MR (n=193), TIM Trio (n=5), and MAGNETOM Vida (n=48) scanners (median delay from MRI to PET scans: 28 days, range: 0-523).

*Neuroimaging Processing*

For all cohorts, T1-weighted MRI scans were processed using FreeSurfer (version 5.3 or 6) and segmented using the Desikan-Killiany atlas ^16^.

In PREVENT-AD, Aβ- and tau-PET scans were preprocessed using a standard pipeline (<https://github.com/villeneuvelab/vlpp>). Briefly, 4D PET images were realigned, averaged, and registered to the corresponding T1 scan, then masked to exclude signal from the scalp and cerebrospinal fluid, and smoothed with a 6mm^3^ Gaussian kernel. Standardized uptake value ratios (SUVRs) for each Desikan-Killaney region were computed by dividing the tracer uptake in the cerebellum grey matter for Aβ-PET scans ^17^, and the inferior cerebellar grey matter for tau-PET scans ^18^.

In HABS, PET images were first averaged. For PiB, this comprised the first 8-minutes post-injection. For FTP, the first and last frames were removed for participants with the longer scan duration, to ensure comparability across subjects. Images were then co-registered to the corresponding T1-weighted MRI using a 6 DoF rigid body registration in SPM12. Distribution volume ratios (DVRs) for PiB, and SUVRs for FTP, were calculated using the cerebellum grey matter.

In AIBL, PET scans were spatially normalised using CapAIBL^19^ for Aβ and CapAIBL PCA for tau^20^. Tau-PET scans were scaled using the cerebellar grey matter as a reference region.

In Knight ADRC, the PET Unified Pipeline (PUP, https://github.com/ysu001/PUP) was used to process all PET scans. SUVRs were calculated from FreeSurfer derived ROIs using the cerebellar gray matter as the reference region ^21,22^.

*A/T/(N) classification*

Cohort-specific thresholds were employed to establish Aβ positivity using previously employed methods. These thresholds were derived from a global amyloid index, comprising the average SUVR/DVR of lateral and medial frontal, cingulate, parietal, and lateral temporal regions. In PREVENT-AD, an SUVR threshold of 1.29 (22.32 Centiloids) was selected for NAV positivity, calculated as the midpoint between a liberal and conservative threshold. The liberal threshold (SUVR 1.20) was equivalent to 15 Centiloids, which is considered the lowest clinically significant cut-off value for Aβ positivity ^23^. The conservative estimate (SUVR 1.37, 29.13 Centiloids) was derived using 2-component gaussian mixture modeling ^17^. In HABS, a PiB DVR threshold of 1.19 (23.9 Centiloids) was employed ^23,24^. In AIBL, a threshold of 25 centiloids was used. In Knight ADRC, thresholds of 27.1 and 21.9 Centiloids were used for PiB and florbetapir, respectively.

In all cohorts, a temporal meta-ROI was used as the primary measure of tau positivity, comprising the average SUVR of the bilateral entorhinal cortex, amygdala, fusiform gyrus, and inferior and middle temporal gyri^25^. Participants were classified as tau positive if their temporal meta-ROI uptake surpassed 2 standard deviations from the mean uptake of the cognitively unimpaired (at baseline) Aβ- participants in the respective cohort. This produced an SUVR cut-off of 1.30 in the PREVENT-AD, 1.31 in HABS, 1.32 in AIBL, and 1.28 in Knight ADRC. In sensitivity analyses, results using a temporal meta-ROI were compared to those using other regions to classify tau positivity (entorhinal cortex; inferior temporal; or any positive region out of entorhinal, inferior temporal, and meta-ROI; Supplementary Results).

Two measures were employed to determine neurodegeneration positivity: average cortical thickness in a temporal meta-ROI comprising entorhinal cortex, fusiform, inferior temporal, and middle temporal gyri (reported in the main text) ^26^, and bilateral hippocampal volume (summed across hemispheres and reported as a percentage of total intracranial volume; reported in Supplementary Results). Participants were classified as neurodegeneration positive if these measures were below the 20^th^ percentile of A-T- non-progressor participants within the respective cohorts.

*Outcomes*

*MCI classification*

In PREVENT-AD, MCI classification was based on a consensus review committee comprising dementia specialist neuropsychologists and/or physician and expert research staff. This team was blind to any biological biomarker, including PET, MRI and APOE genotype, that could have influenced the classification. Participants performing less than 1.0 standard deviations from demographically-stratified norms on at least two neuropsychological tests (including the RBANS and other measures) were discussed at consensus meetings. Participants with major cognitive complaints, or for whom doubt about cognitive status was raised by the research assistant performing the neuropsychological evaluation, were also discussed. Classification was based on longitudinal cognitive trajectories and relevant clinical history.

In HABS, MCI classification was determined by clinical consensus meetings comprising neuropsychologists, neurologists, and psychiatrists, blind to imaging and genetic biomarkers ^27^. Participants with a global Clinical Dementia Rating (CDR)^28^ score of 0.5 or greater, or performance less than 1.5 standard deviations from the sample mean on the PACC, were brought to consensus.

In AIBL, MCI classification was made by clinical review panels comprising psychiatrists, a neurologist, a geriatrician, and neuropsychologists, blind to imaging and genetic biomarkers ^4^. Participants were flagged for discussion at review if demonstrated any of the following: scored <28/30 on the MMSE, failure on the Logical Memory test (based on the Alzheimer’s Disease Neuroimaging Initiative (ADNI) criteria), other evidence of difficulty on neuropsychological testing, a CDR score of 0.5 or greater, or a medical/personal/informant history suggestive of impaired cognitive function.

In Knight ADRC, MCI classification was based on a CDR score of 0.5 or greater. Raters were
blind to imaging and genetic biomarkers.

*Statistical analyses*

We first compared demographic variables, APOE4 status, MRI measures, and baseline cognitive performance between the AT biomarker groups (A+T+, A+T-, A-T-) using one-way analyses of variance with Tukey’s post hoc tests for continuous variables, and Fisher’s exact tests for categorical variables. We next compared the proportion of MCI progressors versus non-progressors in each of the AT biomarker groups using Fisher’s exact tests, as well as between the A+T+N+ and A+T+N- groups. Cox proportional hazard models were then used to test the risk of MCI progression over time in the A+T+ group relative to the other PET biomarker groups. Demographic (age, sex, education) and clinical (APOE4 status, MMSE performance at time of tau-PET) variables were included as covariates in the Cox models, with data censored at the date of the last clinical follow-up visit or at clinical progression for each participant. In follow-up analyses, continuous measures of neurodegeneration (hippocampal volume/temporal cortical thickness) were also added to the Cox models. We also compared the performance of each of these models to baseline models comprising just the demographic and clinical variables, to determine the additional predictive value of the A, T, and N biomarkers. Finally, we employed linear mixed-effects models to investigate longitudinal cognitive decline across the different AT biomarker groups. Models included random slopes and intercepts for each subject and covariates of age, sex, and education, with the interaction between time (years since cognitive baseline visit) and biomarker group determining differences in cognitive change over time between the groups. To examine whether those individuals not yet progressing to MCI were likely on the clinical pathway, these participants were further divided into cognitively ‘stable’ versus ‘decliners’ based on individual longitudinal cognitive slopes. The proportion of cognitive decliners versus cognitively stable in each biomarker group were then compared using Fisher’s exact tests.

**Supplementary Results**

*Demographic and biological characteristics across biomarker groups*

Characteristics of participants across biomarker groups are presented in Table 1 in the main text, and in Supplementary Tables 1 and 2 separated by MCI progression status. In PREVENT-AD the A+T+ group was older than the other groups (F(2, 124) = 5.03, *p* = .008; *post hoc p* values < .05). The A-T- group had a lower proportion of APOE4 carriers compared with the other groups (Fisher’s exact *p* values *≤* .008). The A+T- group had thicker temporal cortices than the A+T+ and A-T- groups (F(2, 124) = 6.08, *p* = .003; *post hoc p* values < .03), and the A+T+ group displayed a trend toward smaller hippocampal volumes compared with the A+T- group (F(2, 124) = 3.09, *p* = .05; *post hoc p* = .06). A+T+ participants also displayed worse MMSE scores compared with the other groups (F(2, 124) = 5.59, *p* = .005; *post hoc p* values < .03), and worse baseline delayed memory performance compared with A-T- participants (F(2, 124) = 3.64, *p* = .03; *post hoc* *p* = .02). In HABS, the proportion of APOE4 carrier was highest in the A+T+ group, followed by the A+T- then A-T- groups (Fisher’s exact *p* values *<*.04). The A+T+ had thinner temporal cortices F(2, 146) = 5.44, *p* = .005; *post hoc p* = .005) and smaller hippocampal volumes F(2, 146) = 4.42, *p* = .01; *post hoc p* = .03) compared with the A-T- group. The A+T+ group also had lower MMSE scores compared with the other groups (F(2, 146) = 4.01, *p* = .02, *post hoc p* values < .03). In AIBL, the A-T- group was younger than the other groups (F(2, 44) = 7.47, *p* = .002, *post hoc p* values < .03), and A+T+ participants performed worse on the MMSE compared with the other groups F(2, 44) = 15.03, *p* <.001 , *post hoc p* values < .001). In Knight ADRC, the A+T+ group was older than the A-T- group (F(2, 244) = 3.06, *p* = .05, *post hoc p* = .04) and had a higher proportion of females compared with the A-T- group (Fisher’s exact *p* value = .01). The A-T- group had a smaller proportion of APOE4 carriers compared with the A+T+ and A+T- groups (Fisher’s exact *p* values = .001). The A+T+ group had smaller hippocampal volumes compared with the A+T- and A-T- groups (F(2, 244) = 3.67, *p* = .03, *post hoc p* values ≤ .05).

**Supplementary Table 1 Demographic, pathological and clinical characteristics of participants by MCI progression status and biomarker group in PREVENT-AD and HABS**

| **PREVENT-AD** | | | | | | | | |  | **HABS** | | | | | | | |
| --- | --- | --- | --- | --- | --- | --- | --- | --- | --- | --- | --- | --- | --- | --- | --- | --- | --- |
|  | **A+T+**  (n = 13) | | **A+T-**  (n = 31) | | **A-T+**  (n = 1) | | **A-T-**  (n = 83) | |  | **A+T+**  (n = 11) | | **A+T-**  (n = 36) | | **A-T+**  (n = 4) | | **A-T-**  (n = 102) | |
|  | **CU_**  **CU**  (n=5) | **CU_**  **MCI**  (n=8) | **CU_**  **CU**  (n=30) | **CU_**  **MCI**  (n=1) | **CU_**  **CU**  (n=1) | **CU_**  **MCI**  (n=0) | **CU_**  **CU**  (n=75) | **CU_**  **MCI** (n=8) |  | **CU_CU**  **(n = 6)** | **CU_MCI**  **(n = 5)** | **CU_CU**  **(n = 32)** | **CU_MCI**  **(n = 4)** | **CU_CU**  **(n = 4)** | **CU_MCI**  **(n = 0)** | **CU_CU**  **(n = 101)** | **CU_MCI**  **(n = 1)** |
| **Demographics** |  |  |  |  |  |  |  |  |  |  |  |  |  |  |  |  |  |
| Age, years | 73.29 (5.10) | 70.09 (5.07) | 66.74 (4.65) | 65-70 | 60-65 | NA | 66.80 (4.80) | 68.87 (3.73) |  | 78.50 (4.53) | 78.45 (6.49) | 77.19 (6.16) | 79.75 (6.66) | 84.06 (0.37) | NA | 74.97 (6.23) | 80-85 |
| Sex, F:M  (% F) | 5:0 (100) | 5:3 (62.50) | 24:6 (80) | 1:0  (100) | 0:1 (0) | NA | 53:22 (70.67) | 7:1  (87.5) |  | 5:1  (83.33) | 4:1 (80) | 16:16 (50) | 2:2 (50) | 2:2 (50) | NA | 56:45 (55.45) | 1 (100) |
| Education, years | 11.60 (1.52) | 14.63 (2.72) | 15.10 (3.01) | 18.0 | 20 | NA | 15.43 (3.35) | 14.88 (4.02) |  | 16.33 (2.34) | 18 (1.41) | 15.97 (2.89) | 16.75 (2.99) | 18 (1.63) | NA | 15.95 (3.23) | 12.0 |
| APOE ɛ4 carriers, n (%) | 4  (80) | 5 (62.50) | 17 (56.67) | 1  (100) | 0 (0) | NA | 19 (25.33) | 4  (50) |  | 6  (100) | 4  (80) | 18 (56.25) | 2  (50) | 1  (25) | NA | 15 (14.85) | 0  (0) |
| **PET** |  |  |  |  |  |  |  |  |  |  |  |  |  |  |  |  |  |
| Global Aβ Centiloid | 67.80 (27.99) | 80.91 (47.64) | 45.57 (28.49) | 25.22 | 18.33 | NA | 11.91 (5.03) | 14.21 (5.79) |  | 51.62 (13.39) | 54.35 (13.55) | 43.28 (19.23) | 43.33 (14.64) | 10.45 (1.42) | NA | 7.10 (4.07) | 7.64 |
| Temporal meta-ROI SUVR | 1.43 (0.13) | 1.41 (0.21) | 1.18 (0.06) | 1.22 | 1.30 | NA | 1.14 (0.07) | 1.20 (0.05) |  | 1.41 (0.02) | 1.40 (0.08) | 1.20 (0.07) | 1.23 (0.07) | 1.33 (0.01) | NA | 1.16 (0.06) | 1.23 |
| **MRI** |  |  |  |  |  |  |  |  |  |  |  |  |  |  |  |  |  |
| Temporal cortical thickness | 2.82 (0.08) | 2.83 (0.14) | 2.95 (0.09) | 2.86 | 2.91 | NA | 2.89 (0.12) | 2.85 (0.11) |  | 2.86 (0.11) | 2.56 (0.10) | 2.85 (0.16) | 2.70 (0.29) | 2.88 (0.13) | NA | 2.88 (0.14) | 2.73 |
| Hippocampal volume (% of TIV) | 0.54 (0.04) | 0.51 (0.07) | 0.56 (0.06) | 0.59 | 0.56 | NA | 0.54 (0.06) | 0.52 (0.07) |  | 0.49 (0.05) | 0.40 (0.03) | 0.47 (0.06) | 0.48 (0.05) | 0.46 (0.04) | NA | 0.49 (0.06) | 0.48 |
| **Cognition** |  |  |  |  |  |  |  |  |  |  |  |  |  |  |  |  |  |
| MMSE (/30) | 27.80 (1.92) | 27.88 (1.46) | 29.17 (0.79) | 30 | 30 | NA | 28.83 (1.29) | 28.38 (1.30) |  | 28.83 (0.75) | 28.00 (1.58) | 29.34 (0.79) | 29.00 (1.41) | 29.75 (0.50) | NA | 29.36 (0.88) | 25 |
| RBANS, baseline |  |  |  |  |  |  |  |  |  |  |  |  |  |  |  |  |  |
| Global Cognition | -0.82 (1.07) | -0.08 (0.86) | 0.01 (0.90) | -0.15 | 0.32 | NA | 0.01 (0.83) | -1.07 (0.70) |  | NA | NA | NA | NA | NA | NA | NA | NA |
| PACC-5, baseline | NA | NA | NA | NA | NA | NA | NA | NA |  | -0.01 (0.35) | 0.30 (0.73) | 0.08 (0.52) | 0.15 (0.55) | -0.17 (0.53) | NA | 0.12 (0.65) | -0.98 |

Data is presented as mean (standard deviation) unless otherwise specified. Aβ = amyloid βeta; APOE = apolipoprotein E; CU_CU = Cognitively unimpaired at time of PET, remaining cognitively unimpaired during follow-up; CU_MCI = Cognitively unimpaired at time of PET, progressing to MCI during follow-up; meta-ROI = meta region-of-interest; MCI = Mild Cognitive Impairment; MMSE = Mini Mental State Examination; PACC = Preclinical Alzheimer’s Composite Score; PET = Positron Emission Tomography; RBANS = Repeatable Battery for the Assessment of Neuropsychological Status; SUVR = standardized uptake value ratio; TIV = total intracranial volume.

*Notes:* Age and MMSE performance are calculated at the time of tau PET. APOE ɛ4 carriers had at least one copy of the ɛ4 allele. Cognitive variables (RBANS, PACC) represent cohort-derived z-scores.

**Supplementary Table 2 Demographic, pathological and clinical characteristics of participants by MCI progression status and biomarker group in AIBL and Knight ADRC**

| **AIBL** | | | | | | | | |  | **Knight ADRC** | | | | | | | |
| --- | --- | --- | --- | --- | --- | --- | --- | --- | --- | --- | --- | --- | --- | --- | --- | --- | --- |
|  | **A+T+**  (n = 6) | | **A+T-**  (n = 10) | | **A-T+**  (n = 1) | | **A-T-**  (n = 31) | |  | **A+T+**  (n = 19) | | **A+T-**  (n = 57) | | **A-T+**  (n = 4) | | **A-T-**  (n = 171) | |
|  | **CU_**  **CU**  (n=1) | **CU_**  **MCI**  (n=5) | **CU_**  **CU**  (n=8) | **CU_**  **MCI**  (n=2) | **CU_**  **CU**  (n=1) | **CU_**  **MCI**  (n=0) | **CU_**  **CU**  (n=30) | **CU_**  **MCI** (n=1) |  | **CU_CU**  (n=13) | **CU_MCI**  (n=6) | **CU_CU**  (n=54) | **CU_MCI**  (n=3) | **CU_CU**  (n=4) | **CU_MCI**  (n=0) | **CU_CU**  (n=158) | **CU_MCI**  (n=13) |
| **Demographics** |  |  |  |  |  |  |  |  |  |  |  |  |  |  |  |  |  |
| Age, years | 80-85 | 78.2 (6.83) | 79.25 (8.45) | 80.50 (6.36) | 80-85 | NA | 71.77 (5.10) | 80-85 |  | 74.87 (6.05) | 75.36 (5.78) | 71.81 (5.19) | 77.22 (7.53) | 70.79 (4.74) | NA | 71.46 (5.86) | 73.62 (4.66) |
| Sex, F:M  (% F) | 1:0 (100) | 4:1 (80) | 5:3 (62.50) | 1:1  (50) | 1:0 (100) | NA | 17:13 (56.67) | 0:1 (0) |  | 10:3 (76.92) | 5:1 (83.33) | 34:20 (62.96) | 2:1 (66.67) | 3:1 (75) | NA | 78:80 (49.37) | 5:8 (38.46) |
| Education, years | 9 | 9.80 (3.03) | 13.00 (2.62) | 9.00 (0.00) | 15 | NA | 11.87 (2.96) | 15 |  | 16.00 (2.48) | 15.83 (2.04) | 16.65 (2.23) | 16.33 (2.89) | 16.00 (1.63) | NA | 16.35 (2.44) | 15.46 (2.50) |
| APOE ɛ4 carriers, n (%) | 1  (100) | 1 (20) | 2 (25) | 1  (50) | 0 (0) | NA | 7 (23.33) | 0 (0) |  | 8 (61.54) | 5 (83.33) | 25 (46.30) | 1 (33.33) | 0 (0) | NA | 35 (22.15) | 2 (15.38) |
| **PET** |  |  |  |  |  |  |  |  |  |  |  |  |  |  |  |  |  |
| Global Aβ Centiloid | 35.00 | 79.80 (32.48) | 67.88 (24.76) | 49.00 (2.83) | -16 | NA | -5.13 (9.38) | 24 |  | 80.42 (39.65) | 78.50 (34.13) | 52.46 (26.53) | 109.43 (47.03) | 6.67 (7.95) | NA | 3.61 (10.04) | 0.14 (10.57) |
| Temporal meta-ROI SUVR | 1.39 | 1.54 (0.17) | 1.17 (0.11) | 1.24 (0.09) | 1.33 | NA | 1.13 (0.09) | 1.07 |  | 1.35 (0.11) | 1.35 (0.06) | 1.16 (0.07) | 1.18 (0.06) | 1.30 (0.02) | NA | 1.13 (0.07) | 1.13 (0.07) |
| **MRI** |  |  |  |  |  |  |  |  |  |  |  |  |  |  |  |  |  |
| Temporal cortical thickness | 2.88 | 2.80 (0.09) | 2.89 (0.11) | 2.75 (0.01) | 3.02 | NA | 2.92 (0.09) | 2.57 |  | 2.79 (0.15) | 2.85 (0.22) | 2.87 (0.13) | 2.81 (0.13) | 2.82 (0.15) | NA | 2.84 (0.13) | 2.71 (0.13) |
| Hippocampal volume (% of TIV) | 0.48 | 0.51 (0.02) | 0.49 (0.03) | 0.41 (0.01) | 0.57 | NA | 0.52 (0.06) | 0.42 |  | 0.48 (0.07) | 0.46 (0.09) | 0.52 (0.07) | 0.49 (0.08) | 0.51 (0.07) | NA | 0.52 (0.07) | 0.47 (0.09) |
| **Cognition** |  |  |  |  |  |  |  |  |  |  |  |  |  |  |  |  |  |
| MMSE (/30) | 26 | 25.80 (2.28) | 29.00 (1.51) | 28.00 (1.41) | 27 | NA | 28.97 (0.96) | 27 |  | 29.23 (1.01) | 29.00  (1.55) | 29.39 (1.02) | 28.67 (1.53) | 29.00 (1.41) | NA | 29.31 (1.08) | 28.77 (1.09) |
| RBANS, baseline |  |  |  |  |  |  |  |  |  |  |  |  |  |  |  |  |  |
| Global Cognition | NA | NA | NA | NA | NA | NA | 0.01 (0.83) | -1.07 (0.70) |  | NA | NA | NA | NA | NA | NA | NA | NA |
| PACC, baseline | -0.25 | -0.66 (0.63) | -0.52 (0.98) | -1.48 (0.13) | -1.40 | NA | NA | NA |  | 0.00 (0.57) | -0.45 (0.50) | 0.04 (0.59) | 0.05 (0.22) | 0.15 (0.35) | NA | 0.01 (0.75) | -0.10 (0.63) |

Data is presented as mean (standard deviation) unless otherwise specified. Aβ = amyloid βeta; APOE = apolipoprotein E; CU_CU = Cognitively unimpaired at time of PET, remaining cognitively unimpaired during follow-up; CU_MCI = Cognitively unimpaired at time of PET, progressing to MCI during follow-up; meta-ROI = meta region-of-interest; MCI = Mild Cognitive Impairment; MMSE = Mini Mental State Examination; PACC = Preclinical Alzheimer’s Composite Score; PET = Positron Emission Tomography; RBANS = Repeatable Battery for the Assessment of Neuropsychological Status; SUVR = standardized uptake value ratio; TIV = total intracranial volume.

*Notes:* Age and MMSE performance are calculated at the time of tau PET. Education data was collected in ranges in AIBL, with the lower boundary of the range reported here. Years of education are therefore likely underestimated (further details in Supplement). APOE ɛ4 carriers had at least one copy of the ɛ4 allele. Cognitive variables (RBANS, PACC) represent cohort-derived z-scores.

*Clinical progression rates across biomarker groups using different regions to define tau positivity*

MCI progression status by AT biomarker group for each of the tau regions are displayed in Supplementary Table 3 and Supplementary Figure 1. Regardless of the region used to define tau positivity, a greater proportion of A+T+ participants progressed to MCI compared with the other biomarker groups, though this difference did not reach statistical significance for entorhinal cortex in AIBL, or for inferior temporal cortex in HABS or Knight ADRC (Supplementary Table 3). Using the meta-ROI to classify tau positivity detected the highest proportion of MCI progressors in the PREVENT-AD and HABS cohorts, and using the inferior temporal ROI detected the highest proportion of MCI progressors in AIBL and Knight ADRC.

**Supplementary Table 3 MCI progression status within biomarker groups across cohorts, using different tau regions to define positivity**

|  | **Temporal meta-ROI** | | | | | **Entorhinal cortex** | | | | | **Inferior temporal cortex** | | | |  | **Any** | | | |
| --- | --- | --- | --- | --- | --- | --- | --- | --- | --- | --- | --- | --- | --- | --- | --- | --- | --- | --- | --- |
|  | PREVENT-AD | HABS | AIBL | Knight ADRC |  | PREVENT-AD | HABS | AIBL | Knight ADRC |  | PREVENT-AD | HABS | AIBL | Knight ADRC |  | PREVENT-AD | HABS | AIBL | Knight ADRC |
| **A+T+** |  |  |  |  |  |  |  |  |  |  |  |  |  |  |  |  |  |  |  |
| CU_MCI: CU_CU (% CU_MCI) | 8:5 (61.54)^a,b^ | 5:6 (45.45)^a,b^ | 5:1 (83.33)^a,b^ | 6:13 (31.58) ^a,b^ |  | 7:8 (46.67)^a,b^ | 6:8 (42.86)^a,b^ | 1:0 (100) | 6:15 (28.57)^a,b^ |  | 5:6 (45.45)^a,b^ | 4:6 (40)^b^ | 4:0 (100)^a,b^ | 3:7 (30)^b^ |  | 9:8 (52.94)^a,b^ | 6:9 (40)^a,b^ | 5:1 (83.33)^a,b^ | 7:18 (28)^a,b^ |
| **A+T-** |  |  |  |  |  |  |  |  |  |  |  |  |  |  |  |  |  |  |  |
| CU_MCI: CU_CU (% CU_MCI) | 1:30 (3.23) | 4:32 (11.11)^c^ | 2:8 (20) | 3:54 (5.26) |  | 2:27 (6.90) | 3:30 (9.09)^c^ | 6:9 (40)^c^ | 3:52 (5.45) |  | 4:29 (12.12) | 5:32 (13.51)^c^ | 3:9 (25) | 6:60 (9.09) |  | 1:26 (3.70) | 3:29 (9.38)^c^ | 2:8 (20) | 2:49 (3.92) |
| **A-T+** |  |  |  |  |  |  |  |  |  |  |  |  |  |  |  |  |  |  |  |
| CU_MCI: CU_CU (% CU_MCI) | 0:1 (0) | 0:4 (0) | 0:1 (0) | 0:4 (0) |  | 0:1 (0) | 0:2 (0) | 0:1 (0) | 1:6 (14.29) |  | 0:1 (0) | 0:2 (0) | 0:2 (0) | 0:2 (0) |  | 0:2 (0) | 0:5 (0) | 0:3(0) | 1:9 (10) |
| **A-T-** |  |  |  |  |  |  |  |  |  |  |  |  |  |  |  |  |  |  |  |
| CU_MCI: CU_CU (% CU_MCI) | 8:75 (9.64) | 1:101 (0.98) | 1:30 (3.23) | 13:158 (7.60) |  | 8:75 (9.64) | 1:103 (.96) | 1:30 (3.23) | 12:156 (7.14) |  | 8:75 (9.64) | 1:103 (0.96) | 1:29 (3.33) | 13:160 (7.51) |  | 8:82 (9.76) | 1:100 (.99) | 1:28 (3.45) | 12:153(7.27) |

CU_CU = Cognitively unimpaired at time of PET, remaining cognitively unimpaired during follow-up; CU_MCI = Cognitively unimpaired at time of PET, progressing to MCI during follow-up

*Notes:* Biomarker group definition is based on Aβ and tau PET scans. Progression status is based on clinical follow-up data at least 12 months after Aβ and tau PET scans. ‘Any’ refers to any positive region out of temporal meta-ROI, entorhinal cortex, and inferior temporal cortex. ^a^ = significant difference between A+T+ and A+T-, ^b^ = significant difference between A+T+ and A-T-, ^c^ = significant difference between A+T- and A-T- at *p* < .05 using Fisher’s exact tests. The A-T+ group is presented for completion but was not included in statistical analysis.

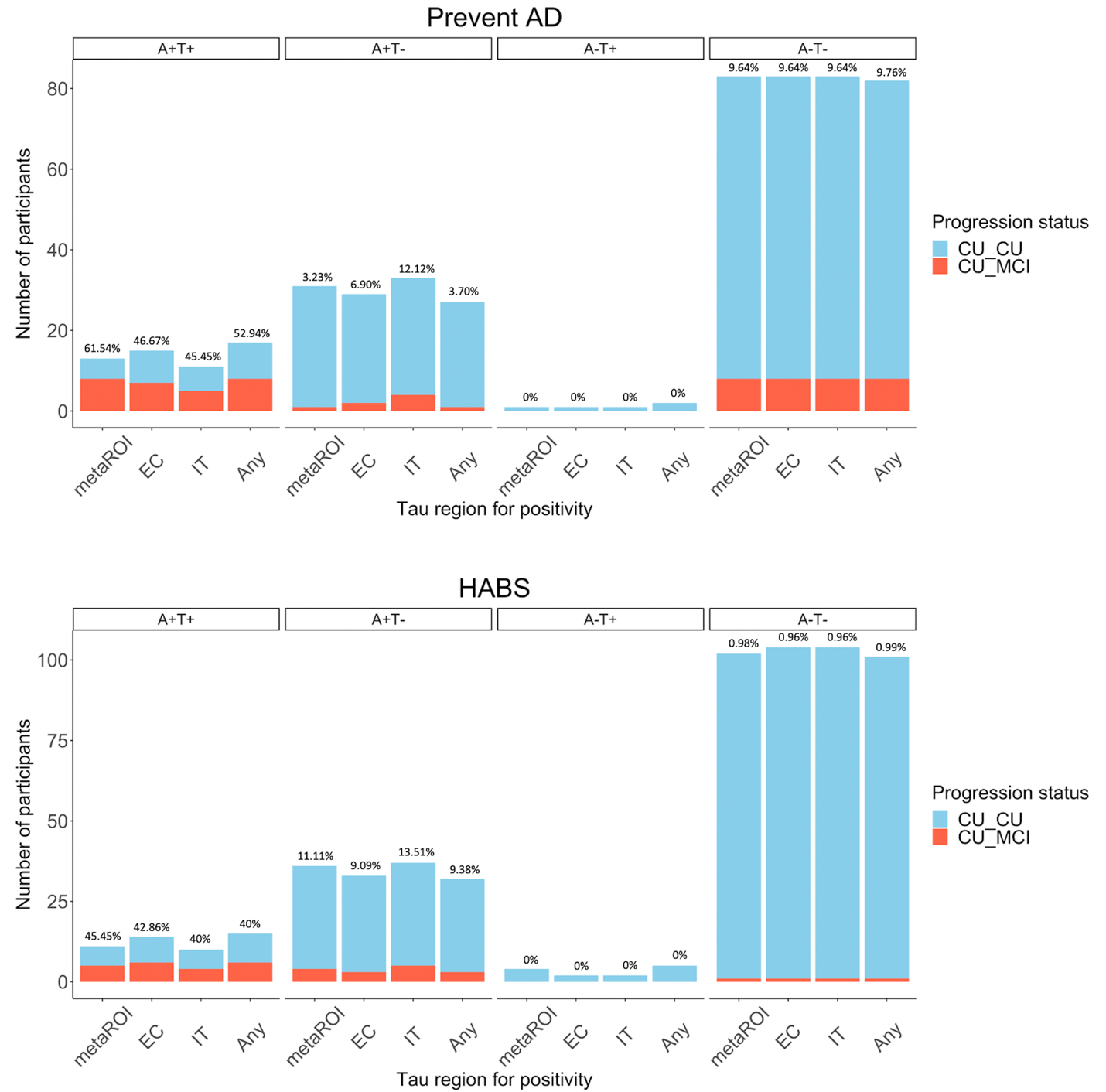

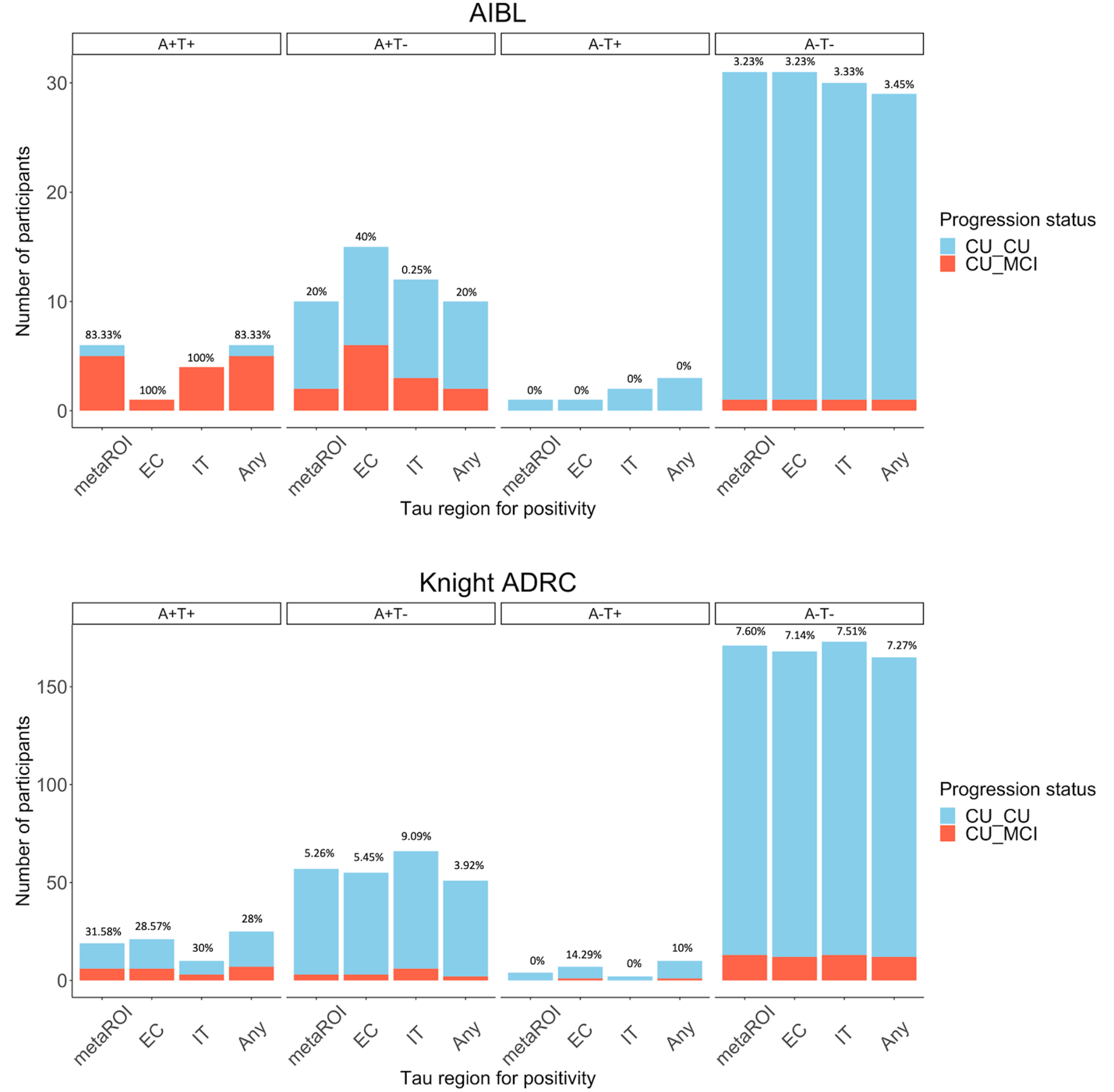
*Supplementary Figure 1.* Number of participants progressing to MCI after PET versus those remaining cognitively unimpaired in each AT biomarker group, across cohorts, using different regions to define tau positivity. Percentage values represent the proportion of MCI progressors within the group. CU_CU = Cognitively unimpaired at time of PET, remaining cognitively unimpaired during follow-up; CU_MCI = Cognitively unimpaired at time of PET, progressing to MCI during follow-up. Any = any positive region out of temporal meta-ROI; entorhinal cortex, and inferior temporal cortex; CU = Cognitively unimpaired at time of PET, remaining cognitively unimpaired during follow-up; EC = entorhinal cortex; IT = inferior temporal cortex; meta-ROI = temporal meta-ROI. *Note:* The A-T+ group is displayed for visualisation purposes but was not included in statistical analysis.

*Clinical progression rates across AT(N) groups using different regions to define neurodegeneration*

Using temporal cortical thickness, evidence of neurodegeneration (N+) in the A+T+ group was associated with increased progression to MCI in HABS (A+T+N+ = 100% (4/4), A+T+N- = 14.29% (1/7)), Fisher’s exact *p* = .02), but not the other cohorts (PREVENT-AD: A+T+N+ = 57.14% (4/7), A+T+N- = 66.67% (4/6), Fisher’s exact *p* = 1.0; AIBL: A+T+N+ = 100% (4/4), A+T+N- = 50% (1/2), Fisher’s exact *p* = .33; Knight ADRC: A+T+N+ = 37.50 % (3/8), A+T+N- = 27.27% (3/11), Fisher’s exact *p* = 1.0). Using hippocampal volume, evidence of neurodegeneration (N+) in the A+T+ group was not associated with increased progression to MCI in any of the cohorts (PREVENT-AD: A+T+N+ = 80% (4/5), A+T+N- = 50% (4/8), Fisher’s exact *p* = .56; HABS: A+T+N+ = 71.43% (5/7), A+T+N- = 0% (0/4), Fisher’s exact *p* = .06; AIBL: A+T+N+ = 66.67% (2/3), A+T+N- = 100% (3/3), Fisher’s exact *p* = 1.0; Knight ADRC: A+T+N+ = 33.33% (3/9), A+T+N- = 30% (3/10), Fisher’s exact *p* = 1.0) (Figure 1B; Supplementary Figure 2).

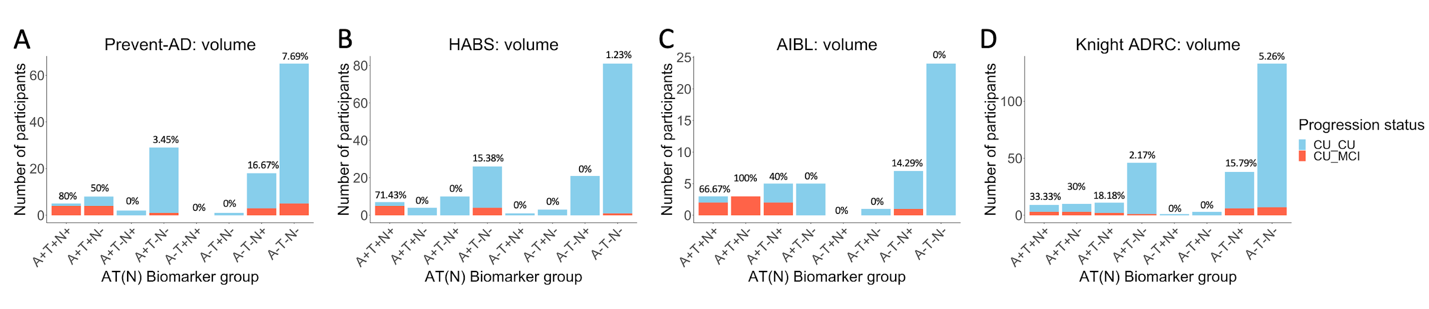
*Supplementary Figure 2.* Number of participants progressing to MCI after PET versus those remaining cognitively unimpaired in each AT(N) biomarker group, using hippocampal volume to define N+, across cohorts. Percentage values represent the proportion of MCI progressors within the group. CU_CU = Cognitively unimpaired at time of PET, remaining cognitively unimpaired during follow-up; CU_MCI = Cognitively unimpaired at time of PET, progressing to MCI during follow-up.

*Effect of biomarker group on on probability of clinical progression across time* *using all data for HABS*

Given shorter follow-ups in the A+T+ group compared with the other biomarker groups in HABS, for the main Cox regression analysis data for all participants in this cohort was censored at the last available time point within the A+T+ group (i.e., 2.59 years). Cox regression statistics and survival curves for models including all data from HABS to predict time to MCI classification for each AT biomarker group are displayed in Supplementary Table 4 and Supplementary Figure 3.

**Supplementary Table 4 Cox proportional hazard models predicting time to incident MCI classification using PET-biomarker groups, using all data in HABS**

|  | HABS | |
| --- | --- | --- |
| *Likelihood ratio* | 35.83 | ***p* < .001** |
| *Concordance* | 0.96 | SE = 0.02 |
| *Variable* | *HR* | *P-value* |
| Biomarker group |  |  |
| A+T- | 68.83 | **<.001** |
| A-T- | 1544.72 | **<.001** |
| Age, years | 0.93 | .23 |
| Sex, M | 0.33 | .18 |
| Education, years | 1.07 | .59 |
| APOE4 carrier | 4.67 | **.03** |

HR = hazard ratio; M = male; SE = standard error.

*Notes:* Hazard ratios for the PET biomarker groups are in reference to the A+T+ group. T+ is defined using the temporal meta-ROI. Inverted hazard ratios are reported for ease of interpretation (i.e., reflecting risk of progression to MCI rather than survival i.e., non-progression). Bold values represent statistical significance at *p* < .05.

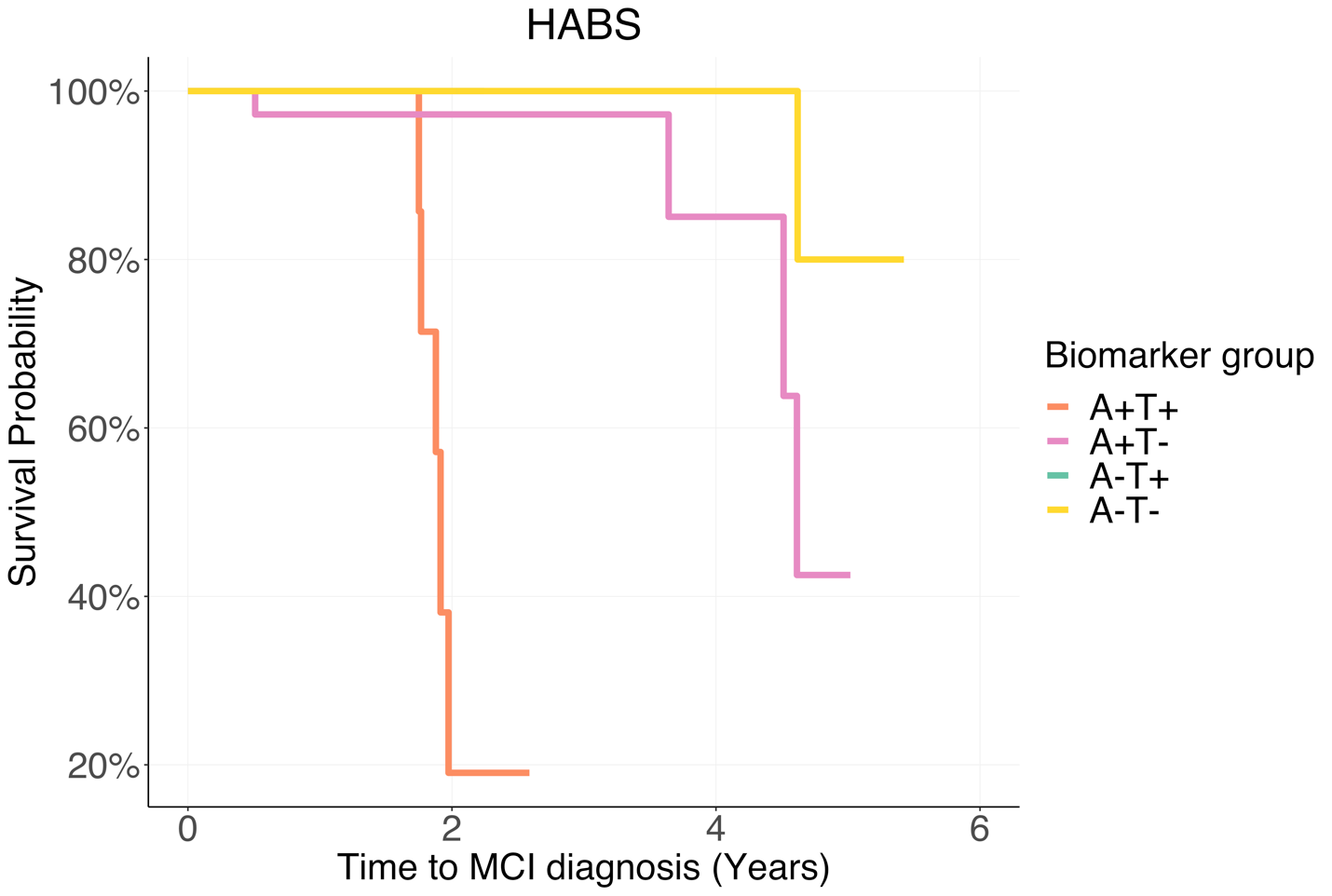

*Supplementary Figure 3.* Survival curves reflecting time from PET scan to MCI for the four PET biomarker groups, in the HABS cohort, using the temporal meta-ROI to define T+ and including data from all available time points. MCI = mild cognitive impairment. *Note:* The A-T+ group is displayed for visualisation purposes but was not included in statistical analyses due to the small sample size.

*Effect of biomarker group on probability of clinical progression across time* *using different regions to define tau positivity*

Cox regression statistics and survival curves representing time to MCI classification for each AT biomarker group, across the different regions to define tau positivity, are displayed in Supplementary Table 5 and Supplementary Figure 4.

**Supplementary Table 5 Cox proportional hazard models predicting time to incident MCI classification using PET-biomarker groups**

|  | **Temporal meta-ROI** | | | | | | | |  | **Entorhinal cortex** | | | | | | | |  | **Inferior temporal cortex** | | | | | | | |  | **Any** | | | | | | | |
| --- | --- | --- | --- | --- | --- | --- | --- | --- | --- | --- | --- | --- | --- | --- | --- | --- | --- | --- | --- | --- | --- | --- | --- | --- | --- | --- | --- | --- | --- | --- | --- | --- | --- | --- | --- |
|  | PREVENT-AD | | HABS | | AIBL | | Knight ADRC | |  | PREVENT-AD | | HABS | | AIBL | | Knight ADRC | |  | PREVENT-AD | | HABS | | AIBL | | Knight ADRC | |  | PREVENT-AD | | HABS | | AIBL | | Knight ADRC | |
| *Likelihood ratio* | 19.37 | *p* = **.004** | 35.06 | *p* < **.001** | 19.37 | *p* = **.004** | 20.57 | *p* = **.002** |  | 16.69 | *p* = **.01** | 30.92 | *p* < **.001** | 21.20 | *p* < **.001** | 15.02 | *p* = **.02** |  | 8.47 | *p* = .20 | 31.16 | *p* < **.001** | 28.57 | *p* < **.001** | 14.03 | *p* = **.03** |  | 15.18 | *p* = **.02** | 30.21 | *p* < **.001** | 18.74 | *p* = **.005** | 16.49 | *p* = **.01** |
| *Concordance* | 0.75 | SE = 0.07 | 0.94 | SE = 0.02 | 0.93 | SE = 0.03 | 0.78 | SE = 0.05 |  | 0.78 | SE = 0.05 | 0.94 | SE = 0.02 | 0.90 | SE = 0.04 | 0.76 | SE = 0.05 |  | 0.73 | SE = 0.05 | 0.93 | SE = 0.03 | 0.93 | SE = 0.05 | 0.76 | SE = 0.04 |  | 0.75 | SE = 0.06 | 0.93 | SE = 0.03 | 0.92 | SE = 0.03 | 0.78 | SE = 0.05 |
| *Variable* | *HR* | *P-value* | *HR* | *P-value* | *HR* | *P-value* | *HR* | *P-value* |  | *HR* | *P-value* | *HR* | *P-value* | *HR* | *P-value* | *HR* | *P-value* |  | *HR* | *P-value* | *HR* | *P-value* | *HR* | *P-value* | *HR* | *P-value* |  | *HR* | *P-value* | *HR* | *P-value* | *HR* | *P-value* | *HR* | *P-value* |
| Biomarker group |  |  |  |  |  |  |  |  |  |  |  |  |  |  |  |  |  |  |  |  |  |  |  |  |  |  |  |  |  |  |  |  |  |  |  |
| A+T- | 22.53 | **.007** | 18.71 | **.007** | 7.98 | **.05** | 10.73 | **.002** |  | 10.52 | **.005** | 5.99 | .07 | 5.98e19 | **<.001** | 4.59 | **.04** |  | 2.81 | .16 | 5.96 | **.05** | 130.63 | **.003** | 5.40 | **.03** |  | 13.37 | **.02** | 4.85 | .10 | 7.94 | **.05** | 6.37 | **.02** |
| A-T- | 5.79 | **.007** | 520.72 | **< .001** | 39.56 | **.004** | 8.20 | **<.001** |  | 3.74 | **.03** | 88.19 | **.002** | NA | NA | 4.64 | **.007** |  | 2.65 | .16 | 145.05 | **.002** | 166.74 | **<.001** | 7.10 | **.003** |  | 3.72 | **.04** | 77.07 | **.003** | 38.16 | **.004** | 4.55 | **.005** |
| Age, years | 1.00 | .81 | 0.91 | .16 | 0.94 | .35 | 0.93 | **.03** |  | 0.93 | .14 | 0.90 | .10 | 0.94 | .32 | 0.92 | **.03** |  | 0.95 | .30 | 0.89 | .07 | 0.76 | **.01** | 0.92 | **.02** |  | 0.98 | .64 | 0.89 | .09 | 0.94 | .35 | 0.93 | **.04** |
| Sex, M | 1.02 | .87 | 0.11 | .03 | 2.68 | .37 | 0.90 | .81 |  | 1.74 | .36 | 0.19 | .08 | 1.21 | .82 | 0.76 | .55 |  | 1.15 | .81 | 0.11 | **.02** | 2.34 | .42 | 0.99 | .98 |  | 1.27 | .68 | 0.16 | **.04** | 2.62 | .38 | 0.77 | .58 |
| Education, years | 0.97 | .52 | 0.98 | .92 | 1.05 | .76 | 1.09 | .36 |  | 0.95 | .49 | 1.01 | .93 | 1.12 | .42 | 1.04 | .70 |  | 0.99 | .93 | 0.91 | .48 | 1.38 | .17 | 1.07 | .45 |  | 0.96 | .64 | 0.98 | .90 | 1.05 | .78 | 1.06 | .53 |
| APOE4 carrier | 0.57 | .33 | 3.48 | .19 | 1.45 | .67 | 0.91 | .85 |  | 0.38 | .08 | 1.69 | .56 | 3.48 | .26 | 1.00 | .99 |  | 0.49 | .18 | 2.13 | .39 | 0.96 | .97 | 0.81 | .66 |  | 0.53 | .25 | 1.83 | .50 | 1.45 | .67 | 0.96 | .93 |

HR = hazard ratio; M = male; SE = standard error.

*Notes:* ‘Any’ refers to any positive region out of temporal meta-ROI, entorhinal cortex and inferior temporal cortex. Hazard ratios for the biomarker groups are in reference to the A+T+ group. Inverted hazard ratios are reported for ease of interpretation (i.e., reflecting risk of progression to MCI rather than ‘survival’ i.e., non-progression). Data for HABS was censored at the last available time point within the A+T+ group, given uneven follow-up times between the biomarker groups. Bold values represent statistical significance at *p* < .05.

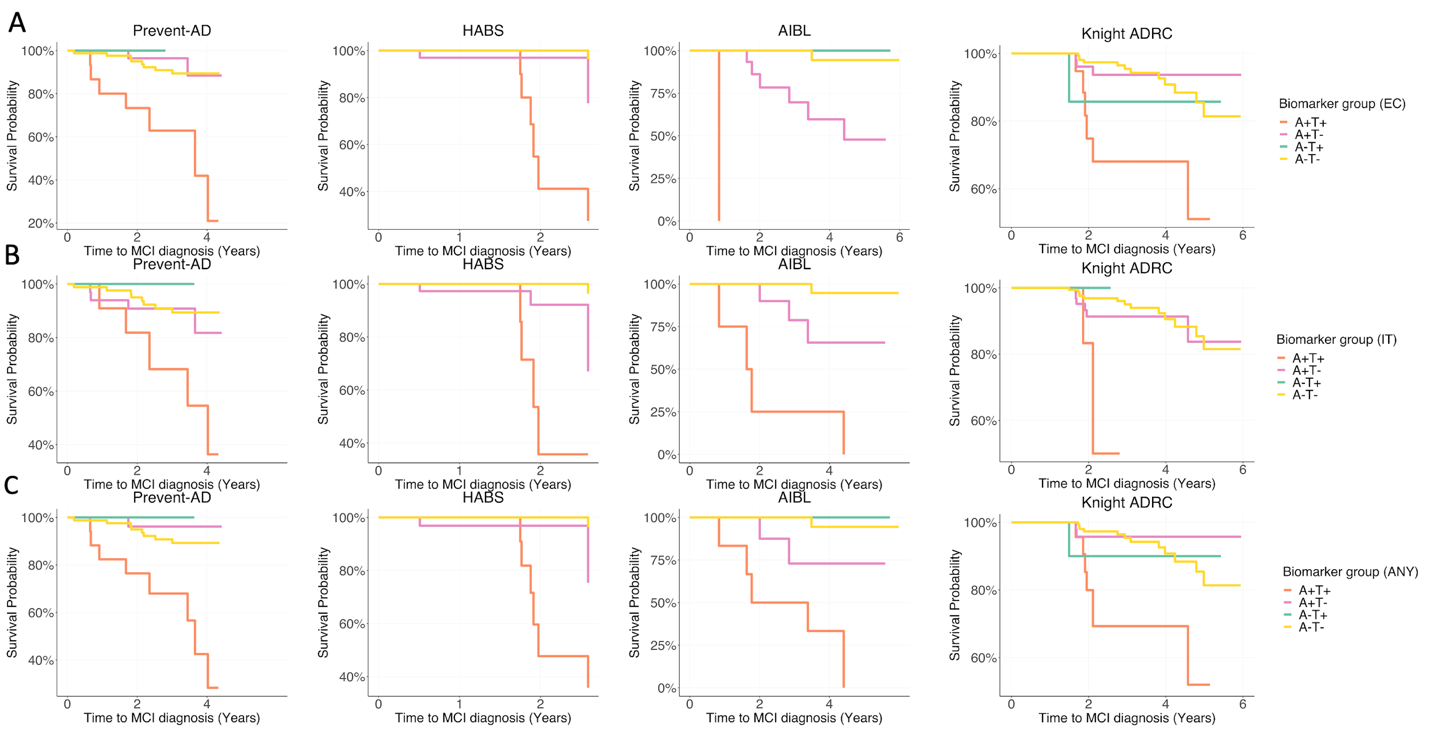

*Supplementary Figure 4.* Survival curves reflecting time from PET scan to MCI for the four PET biomarker groups, across cohorts, using different regions to define tau positivity (A: entorhinal cortex (EC); B: inferior temporal cortex (IT); C: any positive region out of EC, IT, and temporal meta-ROI (ANY)). MCI = mild cognitive impairment. *Note:* The A-T+ group is displayed for visualisation purposes but was not included in statistical analysis. Data for HABS was censored at the last time available point within the A+T+ group, given uneven follow-up times between the biomarker groups.

*Cox models using only demographic/clinical information to predict probability of clinical progression across time*

Cox regression statistics for models using only demographic and clinical information to predict time to MCI classification are displayed in Supplementary Table 6.

**Supplementary Table 6 Cox proportional hazard models predicting time to incident MCI classification using demographic/clinical information**

|  | PREVENT-AD | |  | HABS | |  | AIBL | | Knight ADRC | |
| --- | --- | --- | --- | --- | --- | --- | --- | --- | --- | --- |
| *Likelihood ratio* | 8.51 | *p* = .10 |  | 27.04 | *p* < **.001** |  | 15.01 | *p* = **.01** | 13.81 | *p* = .**02** |
| *Concordance* | 0.72 | SE = 0.06 |  | 0.93 | SE = 0.02 |  | 0.88 | SE = 0.04 | 0.76 | SE = 0.05 |
| *Variable* | *HR* | *P-value* |  | *HR* | *P-value* |  | *HR* | *P-value* | *HR* | *P-value* |
| Age, years | 0.95 | .27 |  | 0.91 | .09 |  | 0.94 | .45 | 0.89 | **.004** |
| Sex, M | 1.04 | .95 |  | 0.27 | .10 |  | 1.12 | .91 | 0.84 | .71 |
| Education, years | 0.96 | .64 |  | 0.67 | **.02** |  | 1.07 | .63 | 0.97 | .79 |
| APOE4 carrier | 0.42 | .08 |  | 0.21 | **.03** |  | 0.81 | .81 | 0.79 | .61 |
| MMSE score | 1.37 | .12 |  | 2.43 | **.003** |  | 1.73 | **.02** | 1.57 | **.01** |

HR = hazard ratio; M = male; SE = standard error.

*Notes:* MMSE score is calculated at the time of tau PET. Inverted hazard ratios are reported for ease of interpretation (i.e., reflecting risk of progression to MCI rather than ‘survival’ i.e., non-progression). Data for HABS was censored at the last available time point within the A+T+ group, given uneven follow-up times between the biomarker groups. Bold values represent statistical significance at *p* < .05.

*Cox models including neurodegenerative measures to predict probability of clinical progression across time*

Cox regression statistics for models incorporating continuous measures of neurodegeneration, in addition to PET biomarker group and demographic/clinical information, to predict time to MCI classification are displayed in Supplementary Table 7.

**Supplementary Table 7 Cox proportional hazard models predicting time to incident MCI classification incorporating continuous measures of neurodegeneration**

|  | **Temporal cortical thickness** | | | | | | | |  | **Hippocampal volume** | | | | | | | |
| --- | --- | --- | --- | --- | --- | --- | --- | --- | --- | --- | --- | --- | --- | --- | --- | --- | --- |
|  | PREVENT-AD | | HABS | | AIBL | | Knight ADRC | |  | PREVENT-AD | | HABS | | AIBL | | Knight ADRC | |
| *Likelihood ratio* | 19.61 | *p* = **.006** | 40.09 | *p* < **.001** | 29.41 | *p* < **.001** | 24.72 | *p* < **.001** |  | 19.71 | *p* = **.006** | 35.87 | *p* < **.001** | 22.02 | *p* = **.003** | 23.24 | *p* = .002 |
| *Concordance* | 0.75 | SE = 0.07 | 0.95 | SE = 0.02 | 0.96 | SE = 0.03 | 0.83 | SE = 0.04 |  | 0.77 | SE = 0.06 | 0.94 | SE = 0.03 | 0.91 | SE = 0.03 | 0.78 | SE = 0.04 |
| *Variable* | *HR* | *P-value* | *HR* | *P-value* | *HR* | *P-value* | *HR* | *P-value* |  | *HR* | *P-value* | *HR* | *P-value* | *HR* | *P-value* | *HR* | *P-value* |
| Biomarker group |  |  |  |  |  |  |  |  |  |  |  |  |  |  |  |  |  |
| A+T- | 20.62 | **.009** | 9.96 | **.04** | 98.44 | .05 | 8.66 | **.005** |  | 19.51 | **.009** | 17.31 | **.01** | 14.73 | **.04** | 8.16 | **.007** |
| A-T- | 5.92 | **.006** | 396.92 | **.004** | 56109.28 | .06 | 7.45 | **<.001** |  | 5.40 | **.009** | 400.76 | **.001** | 93.09 | **.01** | 6.76 | **.002** |
| Age, years | 1.02 | .74 | 0.95 | .40 | 1.48 | .22 | 0.94 | .09 |  | 1.01 | .83 | 0.94 | .39 | 0.95 | .43 | 0.96 | .33 |
| Sex, M | 0.87 | .82 | 0.17 | .08 | 9.00 | .37 | 1.09 | .85 |  | 1.24 | .75 | 0.10 | **.03** | 1.57 | .72 | 1.18 | .73 |
| Education, years | 0.95 | .53 | 0.99 | .97 | 0.69 | .22 | 1.09 | .37 |  | 0.98 | .77 | 1.00 | 1.00 | 0.91 | .64 | 1.09 | .34 |
| APOE4 carrier | 0.58 | .32 | 2.36 | .37 | 0.06 | .15 | 0.89 | .81 |  | 0.57 | .30 | 3.12 | .22 | 1.81 | .52 | 0.87 | .77 |
| Neurodegeneration | 2.27 | .75 | 122.00 | **.03** | 2.23e15 | .11 | 21.91 | **.04** |  | 24.74 | .56 | 1058.15 | .38 | 1.67e11 | .17 | 421.87 | .09 |

*Notes:* Inverted hazard ratios are reported for ease of interpretation (i.e., reflecting risk of progression to MCI rather than ‘survival’ i.e., non-progression). Data for HABS was censored at the last available time point within the A+T+ group, given uneven follow-up times between the biomarker groups. Bold values represent statistical significance at *p* < .05.

*Longitudinal cognition rates across biomarker groups using different regions to define tau positivity*

In all cohorts, A+T+ participants experienced greater longitudinal cognitive decline compared with the other groups regardless of the region used to define tau positivity (*temporal meta-ROI*: PREVENT-AD: β [SE], 0.21 [0.04]; p < 0.001 for A+T-; 0.20 [0.04]; p < 0.001 for A-T-; HABS: β [SE], 0.13 [0.04]; p < 0.001 for A+T-; 0.21 [0.04]; p < 0.001 for A-T-; AIBL: β [SE], 0.37 [0.05]; p < 0.001 for A+T-; 0.40 [0.05]; p < 0.001 for A-T-; Knight ADRC: β [SE], 0.04 [0.02]; p = 0.02 for A+T-; 0.04 [0.02]; p = 0.03 for A-T-; Figure 4A-D; *entorhinal cortex:* PREVENT-AD: β [SE], 0.16 [0.04]; p < 0.001 for A+T-; 0.17 [0.04]; p < 0.001 for A-T-; HABS: β [SE], 0.12 [0.04]; p < 0.001 for A+T-; 0.19 [0.03]; p < 0.001 for A-T-; AIBL: statistics not performed due to only 1 subject in A+T+ group; Knight ADRC: β [SE], 0.04 [0.02]; p = 0.009 for A+T-; 0.03 [0.01]; p = 0.03 for A-T-; *inferior temporal cortex:* PREVENT-AD: β [SE], 0.15 [0.05]; p = 0.002 for A+T-; 0.17 [0.04]; p < 0.001 for A-T-; HABS: β [SE], 0.10 [0.04]; p = 0.01 for A+T-; 0.18 [0.04]; p < 0.001 for A-T-; AIBL: β [SE], 0.40 [0.07]; p < 0.001 for A+T-; 0.46 [0.06]; p < 0.001 for A-T-; Knight ADRC: β [SE], 0.06 [0.02]; p = 0.004 for A+T-; 0.06 [0.02]; p = 0.005 for A-T-; *any:* PREVENT-AD: β [SE], 0.15 [0.04]; p < 0.001 for A+T-; 0.15 [0.04]; p < 0.001 for A-T-; HABS: β [SE], 0.13 [0.03]; p < 0.001 for A+T-; 0.20 [0.03]; p < 0.001 for A-T-; AIBL: β [SE], 0.37 [0.05]; p < 0.001 for A+T-; 0.40 [0.05]; p < 0.001 for A-T-; Knight ADRC: β [SE], 0.04 [0.02]; p = 0.02 for A+T-; 0.03 [0.01]; p = 0.04 for A-T-). Supplementary Figures 5-8 display longitudinal cognitive performance for each AT group (defined using the different tau regions), stratified by MCI progression status for visualisation purposes. Statistical comparisons were not performed between MCI progressors versus non-progressors due to small sample sizes.

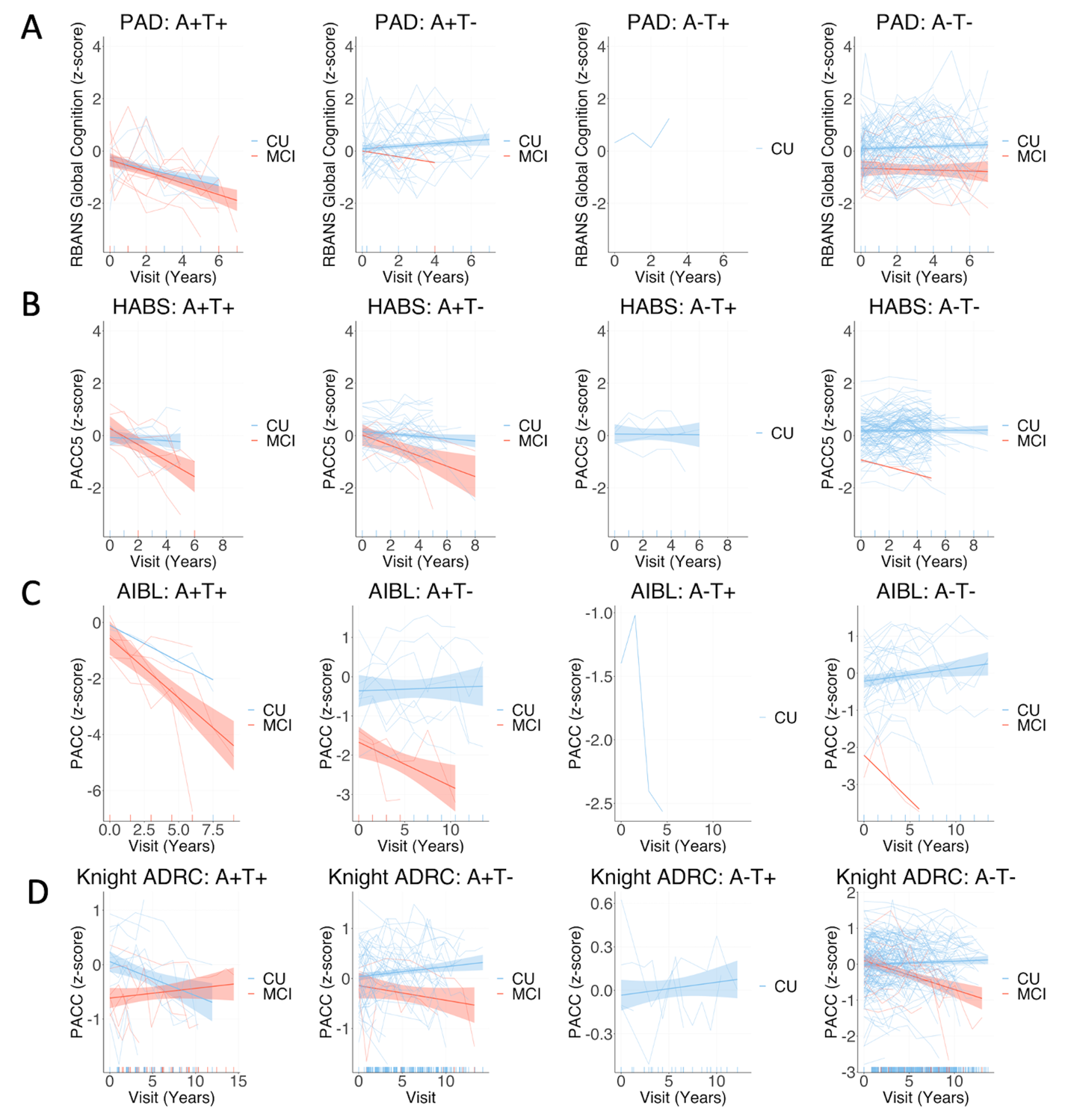

*Supplementary Figure 5.* Group mean and individual longitudinal cognitive slopes for participants who progressed to MCI after PET (red) and those who remained cognitively unimpaired (blue) across each biomarker group across cohorts, using temporal meta-ROI to define tau positivity. Models included random slopes and intercepts for each subject and covariates of age, sex, and years of education*.* CU = Cognitively unimpaired at time of PET, remaining cognitively unimpaired during follow-up; MCI = Cognitively unimpaired at time of PET, progressing to mild cognitive impairment during follow-up. *Notes:* For all cohorts PET was added mid-study, and was therefore performed at different cognitive follow-up visits for each participant. Longitudinal cognition analyses included time points both prior to and after PET scanning. The A-T+ group is displayed for visualisation purposes but was not included in statistical analysis due to the small sample size.

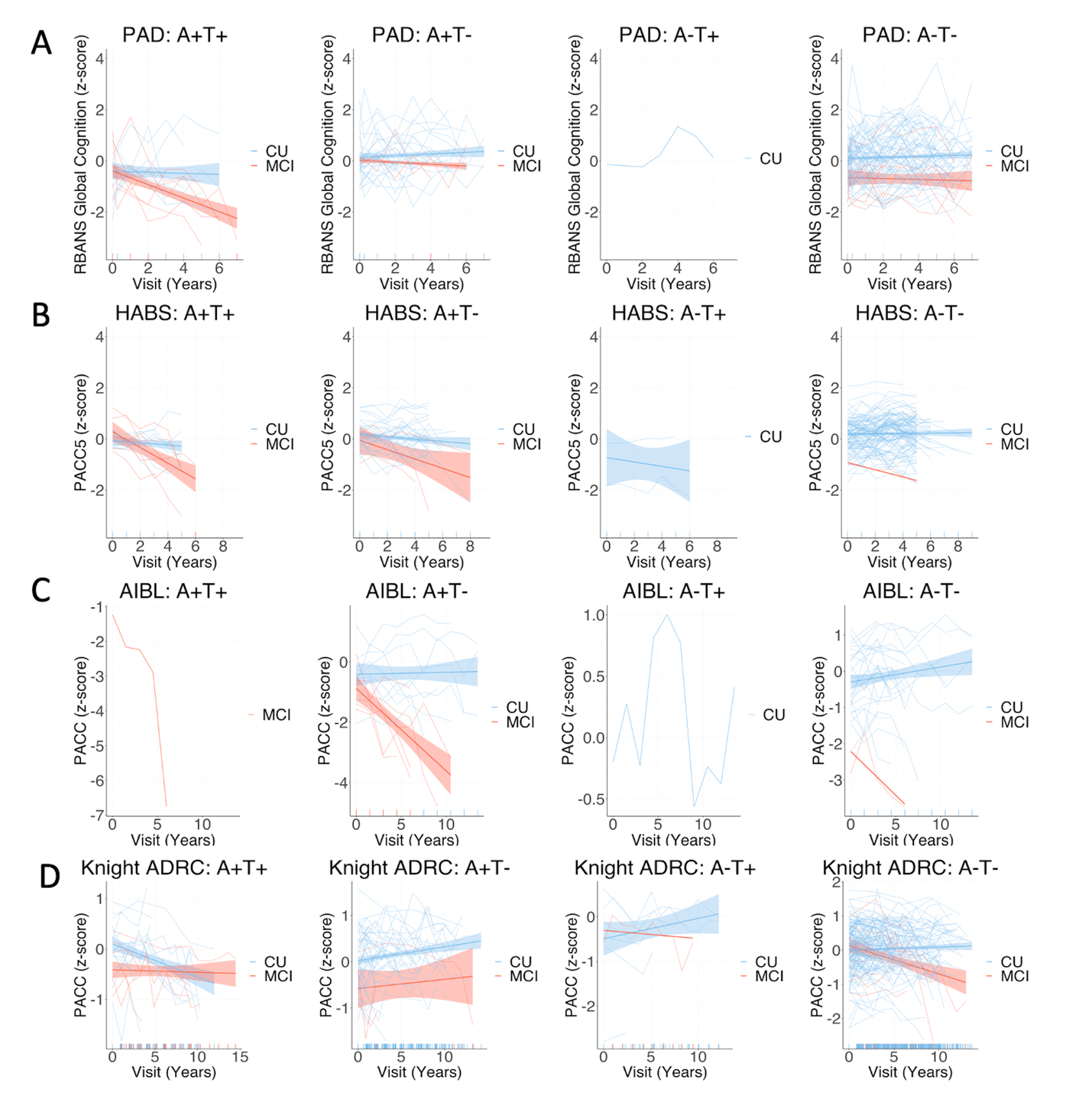

*Supplementary Figure 6.* Group mean and individual longitudinal cognitive slopes for participants who progressed to MCI after PET (red) and those who remained cognitively unimpaired (blue) across each biomarker group across cohorts, using entorhinal cortex to define tau positivity. Models included random slopes and intercepts for each subject and covariates of age, sex, and years of education*.* CU = Cognitively unimpaired at time of PET, remaining cognitively unimpaired during follow-up; MCI = Cognitively unimpaired at time of PET, progressing to mild cognitive impairment during follow-up. *Notes:* For all cohorts PET was added mid-study, and was therefore performed at different cognitive follow-up visits for each participant. Longitudinal cognition analyses included time points both prior to and after PET scanning. The A-T+ group is displayed for visualisation purposes but was not included in statistical analysis due to the small sample size.

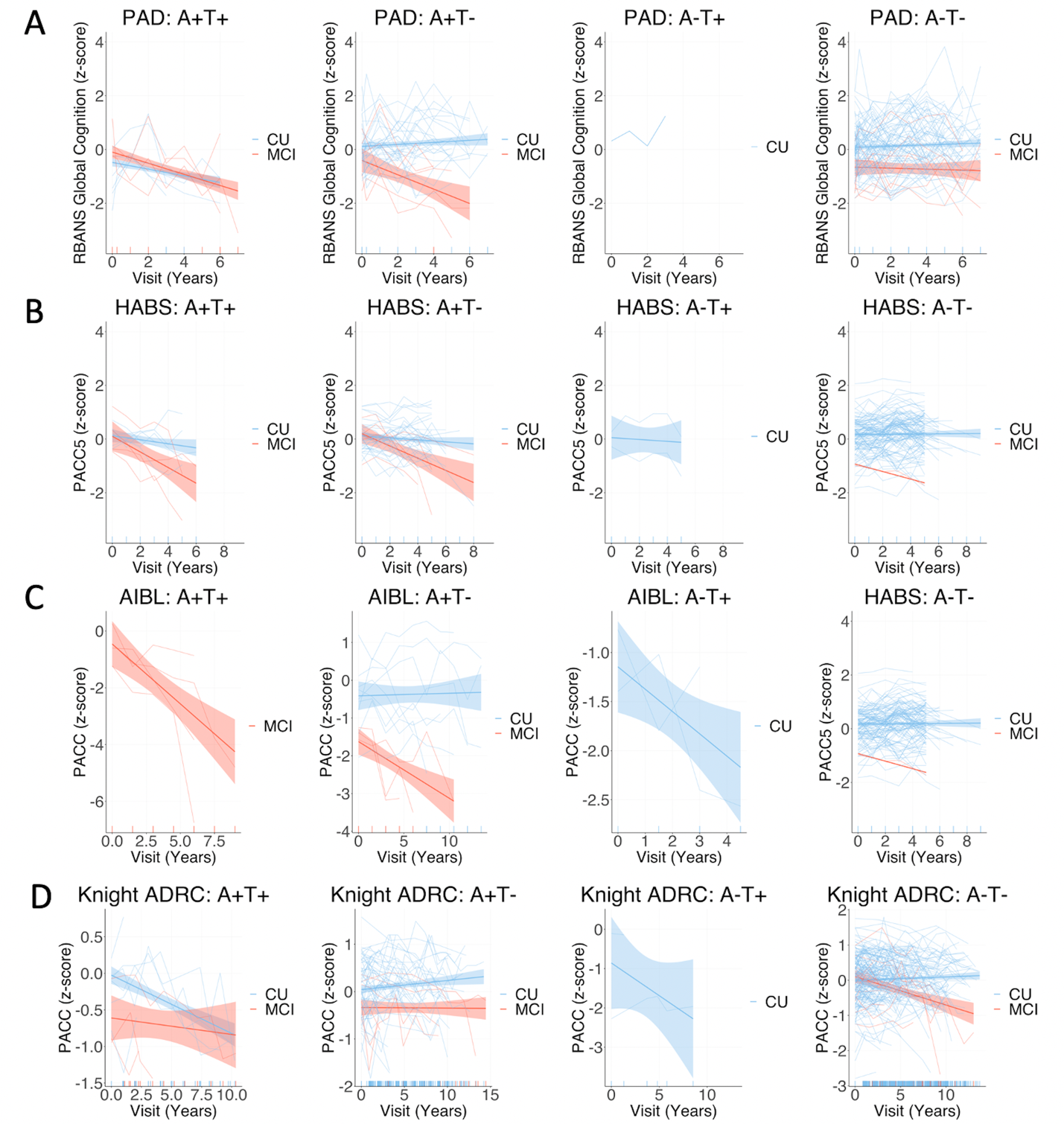

*Supplementary Figure 7.* Group mean and individual longitudinal cognitive slopes for participants who progressed to MCI after PET (red) and those who remained cognitively unimpaired (blue) across each biomarker group across cohorts, using inferior temporal cortex to define tau positivity. Models included random slopes and intercepts for each subject and covariates of age, sex, and years of education*.* CU = Cognitively unimpaired at time of PET, remaining cognitively unimpaired during follow-up; MCI = Cognitively unimpaired at time of PET, progressing to mild cognitive impairment during follow-up. *Notes:* For all cohorts PET was added mid-study, and was therefore performed at different cognitive follow-up visits for each participant. Longitudinal cognition analyses included time points both prior to and after PET scanning. The A-T+ group is displayed for visualisation purposes but was not included in statistical analysis due to the small sample size.

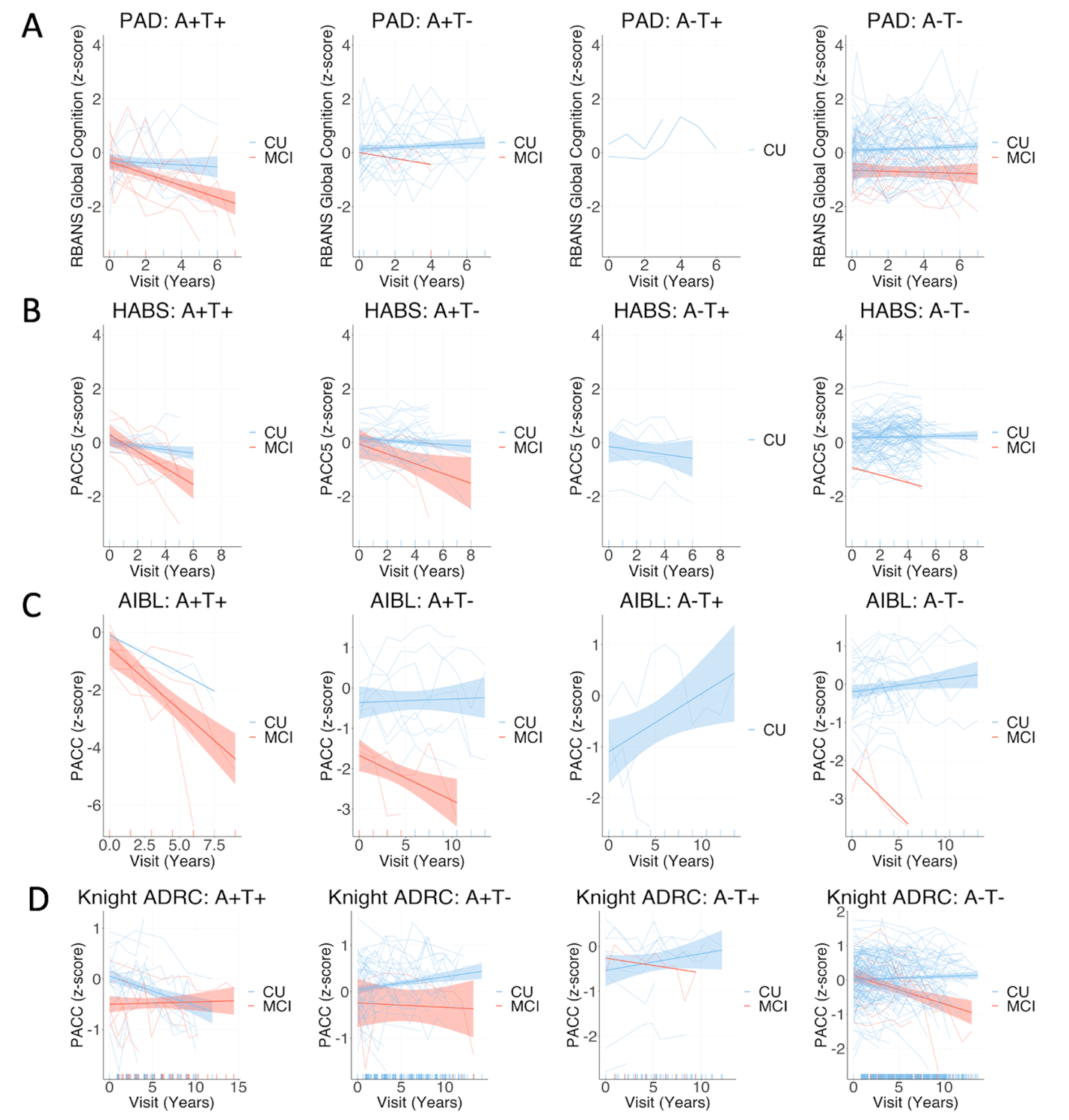

*Supplementary Figure 8.* Group mean and individual longitudinal cognitive slopes for participants who progressed to MCI after PET (red) and those who remained cognitively unimpaired (blue) across each biomarker group across cohorts, using any of temporal meta-ROI, entorhinal cortex, and inferior temporal cortex to define tau positivity. Models included random slopes and intercepts for each subject and covariates of age, sex, and years of education*.* CU = Cognitively unimpaired at time of PET, remaining cognitively unimpaired during follow-up; MCI = Cognitively unimpaired at time of PET, progressing to mild cognitive impairment during follow-up. *Notes:* For all cohorts PET was added mid-study, and was therefore performed at different cognitive follow-up visits for each participant. Longitudinal cognition analyses included time points both prior to and after PET scanning. The A-T+ group is displayed for visualisation purposes but was not included in statistical analysis due to the small sample size.

*Longitudinal cognition for specific cognitive domains in PREVENT-AD*

Supplementary Figure 9 displays longitudinal cognitive performance on each of the five RBANS cognitive index scores in PREVENT-AD, for each PET-biomarker group, stratified by MCI progression status for visualisation purposes. In PREVENT-AD, A+T+ participants experienced greater longitudinal cognitive decline compared with the other groups in immediate (β [SE], 0.31 [0.05]; p < 0.001 for A+T-; 0.28 [0.05]; p < 0.001 for A-T-) and delayed memory (β [SE], 0.27 [0.05]; p < 0.001 for A+T-; 0.22 [0.05]; p < 0.001 for A-T-). For attention and language, greater longitudinal cognitive decline was apparent for the A+T+ compared with A-T- group (attention: β [SE], 0.09 [0.04]; p = 0.03, language: 0.11 [0.05]; p = 0.03) but not the A+T- group (attention: β [SE], 0.07 [0.05]; p = .13; language: 0.10 [0.06]; p = .09). No significant difference was apparent between the A+T+ group and the other groups for visuospatial function (β [SE], 0.05 [0.05]; p = .36 for A+T-; 0.04 [0.05]; p = 0.34 for A-T-).

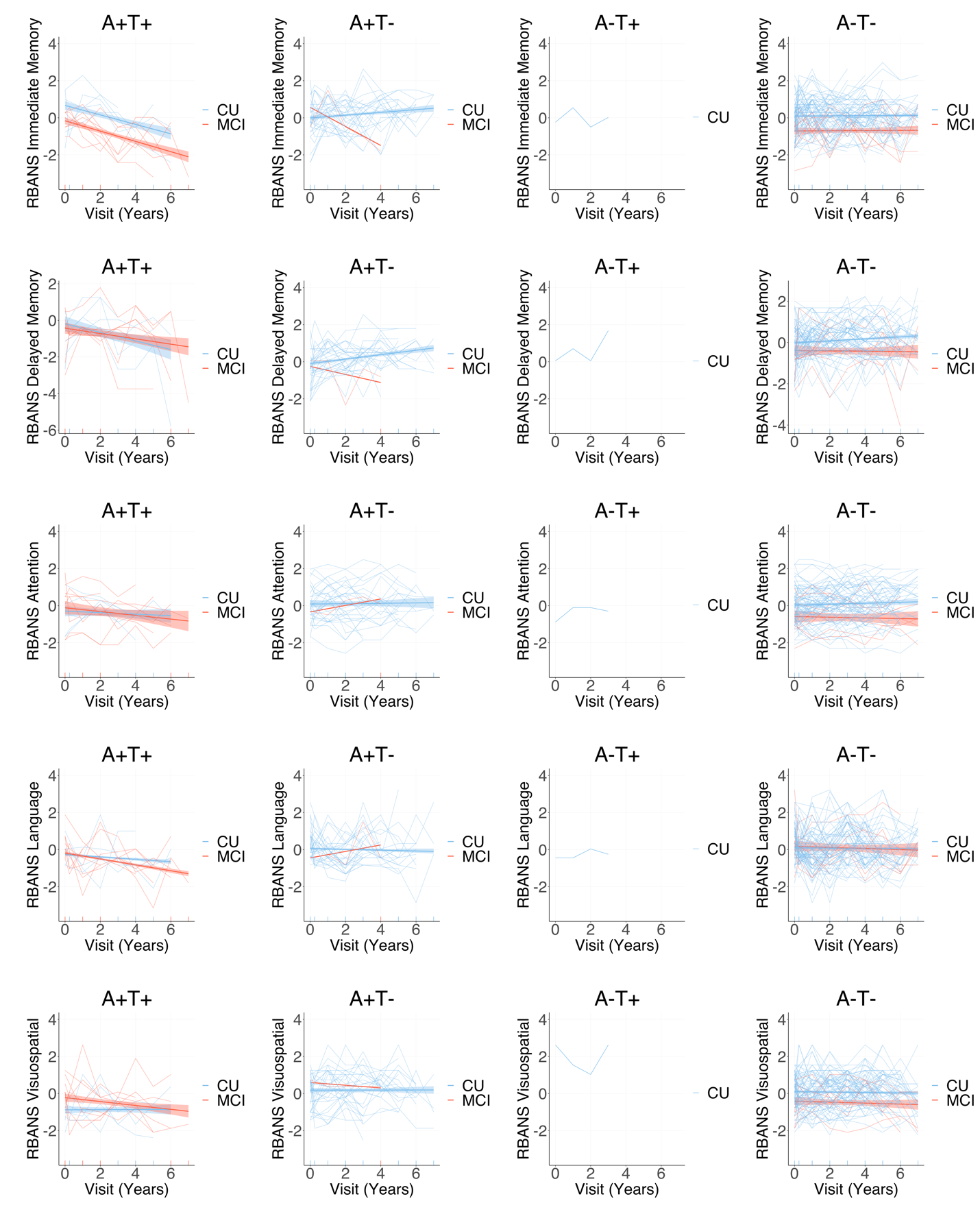

**E**

**D**

**C**

**B**

**A**

*Supplementary Figure 9.* Group mean and individual longitudinal cognitive slopes for each of the RBANS index scores (A: immediate memory; B: delayed memory; C: attention; D: language; E: visuospatial/constructional) for participants who progressed to MCI after PET (red) and those who remained cognitively unimpaired (blue) across each biomarker group in Prevent-AD, using temporal meta-ROI to define tau positivity. Models included random slopes and intercepts for each subject and covariates of age, sex, and years of education*.* CU = Cognitively unimpaired at time of PET, remaining cognitively unimpaired during follow-up; MCI = Cognitively unimpaired at time of PET, progressing to mild cognitive impairment during follow-up. *Notes:* PET was added mid-study, and was therefore performed at different cognitive follow-up visits for each participant. Longitudinal cognition analyses included time points both prior to and after PET scanning. The A-T+ group is displayed for visualisation purposes but was not included in statistical analysis.

*Cognitive decline status of non-progressors across biomarker groups using different regions to define tau positivity*

Cognitive status (declining versus stable) for non-progressors by AT biomarker group, using different regions to define tau positivity, is displayed in Supplementary Figure 10 and Supplementary Table 8. Regardless of the region used to define tau positivity, a greater proportion of A+T+ participants were classified as cognitive decliners, compared with the other biomarker groups, though these differences only reached statistical significance in the PREVENT-AD and Knight ADRC cohorts. The highest proportion of decliners was detected using the inferior temporal cortex to classify tau positivity in PREVENT-AD and Knight ADRC, using ‘any’ region in HABS, and using the temporal meta-ROI in AIBL.

**Supplementary Table 8 Cognitive decline status amongst nonprogressors participants within each PET-biomarker group across cohorts and tau positivity regions**

|  | **Temporal meta-ROI** | | | | **Entorhinal cortex** | | | | **Inferior temporal cortex** | | | | **Any** | | | |
| --- | --- | --- | --- | --- | --- | --- | --- | --- | --- | --- | --- | --- | --- | --- | --- | --- |
|  | PREVENT-AD | HABS | AIBL | Knight ADRC | PREVENT-AD | HABS | AIBL | Knight ADRC | PREVENT-AD | HABS | AIBL | Knight ADRC | PREVENT-AD | HABS | AIBL | Knight ADRC |
| **A+T+** |  |  |  |  |  |  |  |  |  |  |  |  |  |  |  |  |
| CU_Decliner: CU_Stable (% Decliner) | 4:1 (80)^a,b^ | 3:3 (50) | 1:0 (100) | 4:9 (30.77)^a,b^ | 6:2 (75)^a,b^ | 4:4 (50)^b^ | 0:0 (0) | 6:9 (40)^a,b^ | 5:1 (83.33)^a,b^ | 3:3 (50) | 0:0 (0) | 3:4 (42.86)^a,b^ | 6:3 (66.67)^a,b^ | 5:4 (55.56)^b^ | 1:0 (100) | 6:12 (33.33)^a,b^ |
| **A+T-** |  |  |  |  |  |  |  |  |  |  |  |  |  |  |  |  |
| CU_Decliner: CU_Stable (% Decliner) | 3:27 (10) | 11:22(33.33)^c^ | 3:5 (37.5) | 3:51 (5.56) | 1:26 (3.70) | 10:20 (33.33)^c^ | 4:5 (44.44) | 1:51 (1.92) | 2:27 (6.90) | 11:21 (34.38)^c^ | 4:5 (44.44)^c^ | 4:56 (6.67) | 1:25 (3.85) | 9:20 (31.03) | 3:5 (37.5) | 1:48 (2.04) |
| **A-T+** |  |  |  |  |  |  |  |  |  |  |  |  |  |  |  |  |
| CU_Decliner: CU_Stable (% Decliner) | 0:1 (0) | 1:3 (25) | 1:0 (100) | 0:4 (0) | 0:1 (0) | 1:1 (50) | 0:1 (0) | 0:6 (0) | 0:1 (0) | 1:1 (50) | 2:0 (100) | 0:2 (0) | 0:2 (0) | 2:3 (40) | 2:1 (66.67) | 0:9 (0) |
| **A-T-** |  |  |  |  |  |  |  |  |  |  |  |  |  |  |  |  |
| CU_Decliner: CU_Stable (% Decliner) | 12:63 (16) | 15:86 (14.85) | 4:26 (13.33) | 15:143 (9.49) | 12:63 (16) | 15:88 (14.56) | 5:25 (16.67) | 15:141 (9.62) | 12:63 (16) | 15:88 (14.56) | 3:26 (10.34) | 15:145 (9.38) | 12:62 (16.22) | 14:86 (14) | 3:25 (10.71) | 15:138 (9.80) |

CU_Decliner = Cognitively unimpaired participants who did not progress to MCI, but who showed longitudinal cognitive decline; CU_Stable = Cognitively unimpaired participants who did not progress to MCI and remained cognitively stable.

*Notes:* Cognitive decline was characterised by a longitudinal cognitive slope value < 1 SD from the mean of the A-T- non-progressors in each cohort. ‘Any’ refers to any positive region out of temporal meta-ROI, entorhinal cortex, and inferior temporal cortex. ^a^ = significant difference between A+T+ and A+T-, ^b^ = significant difference between A+T+ and A-T-, ^c^ = significant difference between A+T- and A-T- at *p* < .05. The A-T+ group is included for completion but was not included in statistical analysis.

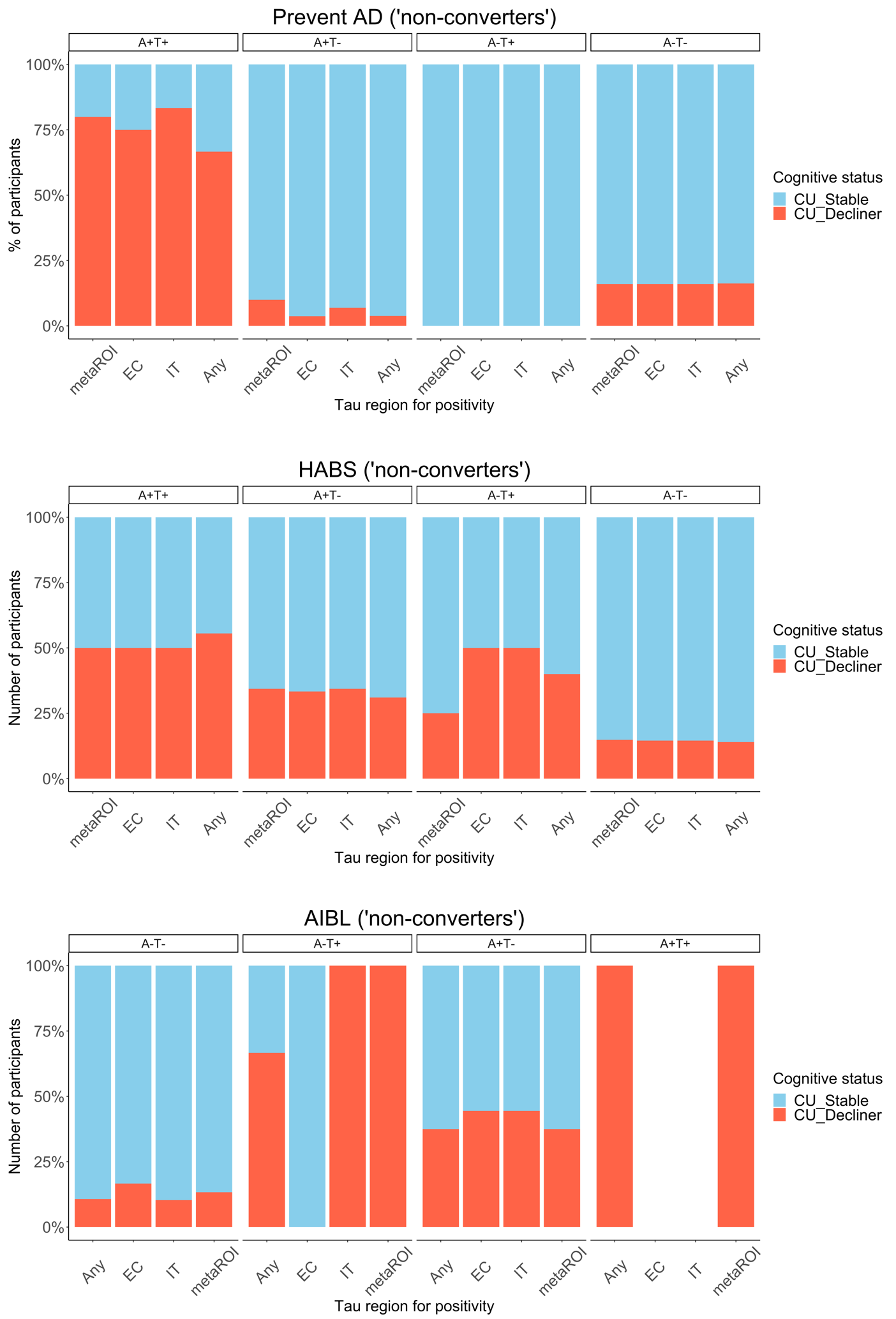

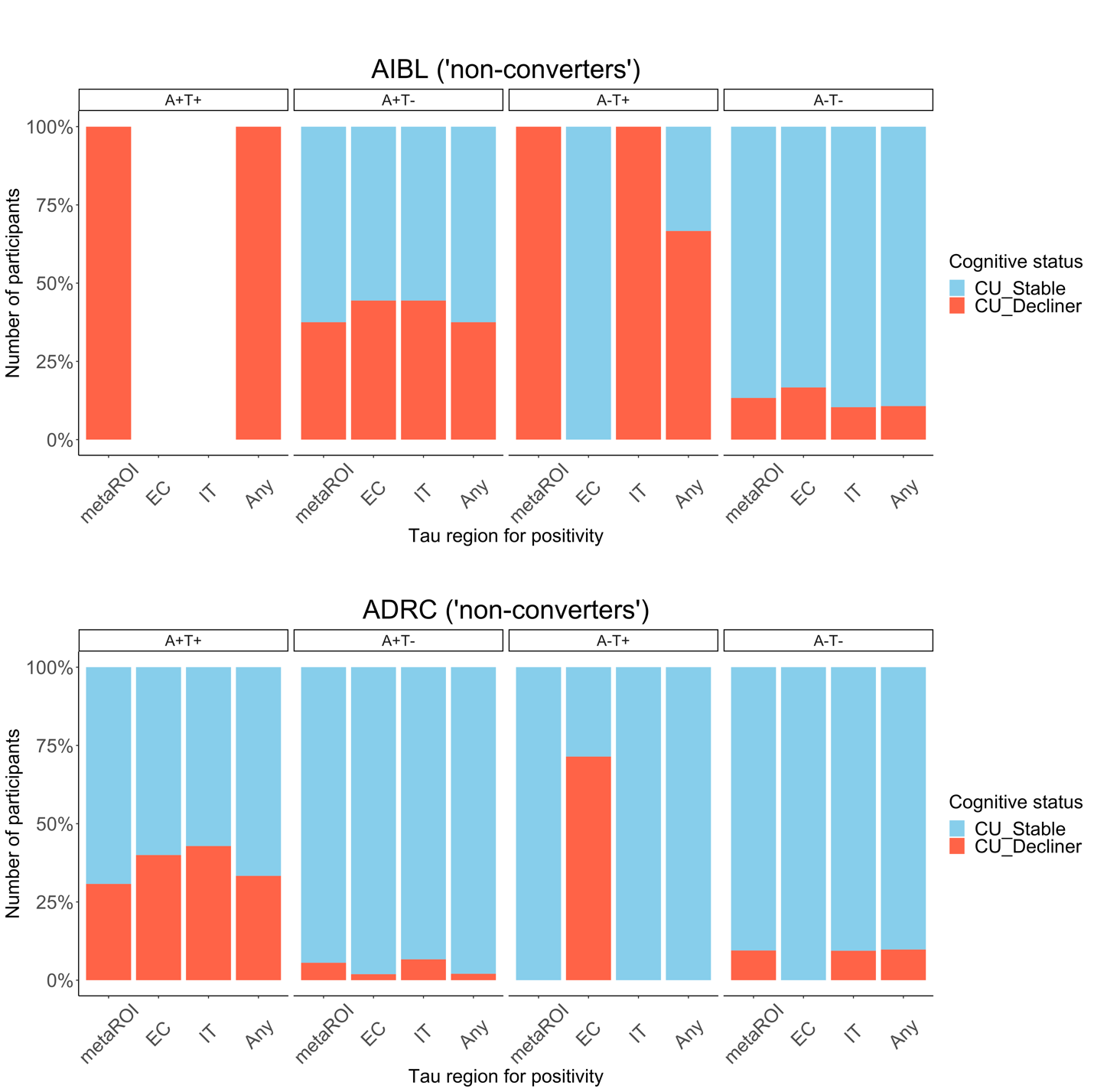

*Supplementary Figure 10.* Percentage of cognitively unimpaired participants (CU), who did not progress to MCI, but who showed longitudinal cognitive decline (CU_Decliner) versus those who remained cognitively stable (CU_Stable) in each AT biomarker group, using different regions to define tau positivity, across cohorts. Any = any positive region out of temporal meta-ROI, entorhinal cortex, and inferior temporal cortex; EC = entorhinal cortex; IT = inferior temporal cortex, meta-ROI = temporal meta-ROI. *Note:* The A-T+ group is displayed for visualisation purposes but was not included in statistical analysis.
